# IL-13 is a driver of COVID-19 severity

**DOI:** 10.1101/2020.06.18.20134353

**Authors:** Alexandra N. Donlan, Tara E. Sutherland, Chelsea Marie, Saskia Preissner, Ben T. Bradley, Rebecca M. Carpenter, Jeffrey M. Sturek, Jennie Z. Ma, G. Brett Moreau, Jeffrey R. Donowitz, Gregory A. Buck, Myrna G. Serrano, Stacey L. Burgess, Mayuresh M. Abhyankar, Cameron Mura, Philip E. Bourne, Robert Preissner, Mary K. Young, Genevieve R. Lyons, Johanna J. Loomba, Sarah J Ratcliffe, Melinda D. Poulter, Amy J. Mathers, Anthony Day, Barbara J. Mann, Judith E. Allen, William A. Petri

**Affiliations:** Division of Infectious Diseases and International Health, Department of Medicine, University of Virginia School of Medicine, Charlottesville VA 22908 USA; Department of Microbiology, Immunology and Cancer Biology, University of Virginia School of Medicine, Charlottesville VA 22908 USA; Lydia Becker Institute of Immunology and Inflammation, School of Biological Sciences, University of Manchester, Manchester Academic Health Sciences Centre, Manchester M13 9PL, United Kingdom; Department Oral and Maxillofacial Surgery, Charité – Universita□tsmedizin Berlin, Freie Universita□t Berlin, Humboldt-Universita□t zu Berlin, and Berlin Institute of Health, Augustenburger Platz 1, 13353 Berlin, Germany; Department of Laboratory Medicine and Pathology, University of Washington, Seattle WA 98109 USA; Division of Pulmonary and Critical Care Medicine, Department of Medicine, University of Virginia School of Medicine, Charlottesville VA 22908 USA; Department of Public Health Sciences, University of Virginia School of Medicine, Charlottesville VA 22908 USA; Division of Pediatric Infectious Diseases, Children’s Hospital of Richmond, Virginia Commonwealth University, Richmond VA 23298 USA; Department of Microbiology and Immunology, School of Medicine, Virginia Commonwealth University, Richmond VA 23298 USA; School of Data Science and Department of Biomedical Engineering University of Virginia, Charlottesville, VA 22904; Science-IT and Institute of Physiology, Charité–Universita□tsmedizin Berlin, corporate member of Freie Universita□t Berlin, Humboldt-Universita□t zu Berlin, and Berlin Institute of Health, Philippstrasse 12, 10115 Berlin, Germany; Integrated Translational Health Research Institute (iTHRIV), University of Virginia School of Medicine, Charlottesville VA 22908 USA; Department of Pathology University of Virginia School of Medicine, Charlottesville VA 22908 USA; Wellcome Trust Centre for Cell-Matrix Research, School of Biological Sciences, Faculty of Biology Medicine and Health, University of Manchester, Manchester Academic Health Sciences Centre, Manchester M13 9PT, United Kingdom

## Abstract

Immune dysregulation is characteristic of the more severe stages of SARS-CoV-2 infection. Understanding the mechanisms by which the immune system contributes to COVID-19 severity may open new avenues to treatment. Here we report that elevated interleukin-13 (IL-13) was associated with the need for mechanical ventilation in two independent patient cohorts. In addition, patients who acquired COVID-19 while prescribed Dupilumab had less severe disease. In SARS-CoV-2 infected mice, IL-13 neutralization reduced death and disease severity without affecting viral load, demonstrating an immunopathogenic role for this cytokine. Following anti-IL-13 treatment in infected mice, in the lung, hyaluronan synthase 1 (*Has1*) was the most downregulated gene and hyaluronan accumulation was decreased. Blockade of the hyaluronan receptor, CD44, reduced mortality in infected mice, supporting the importance of hyaluronan as a pathogenic mediator, and indicating a new role for IL-13 in lung disease. Understanding the role of IL-13 and hyaluronan has important implications for therapy of COVID-19 and potentially other pulmonary diseases.

**Summary:** L-13 levels are elevated in patients with severe COVID-19. In a mouse model of disease, IL-13 neutralization results in reduced disease and lung hyaluronan deposition. Similarly, blockade of hyaluronan’s receptor, CD44, reduces disease, highlighting a novel mechanism for IL-13-mediated pathology.

## Main Text

SARS-CoV-2, the infectious agent causing the ongoing global COVID-19 pandemic, is a virus that primarily infects the lower respiratory tract of hosts by gaining entry to cells via the receptor angiotensin converting enzyme 2 (ACE2) facilitated by transmembrane receptor neuropilin-1 (Cantuti-Castelvetri et al., 2020; Hoffmann et al., 2020). The clinical course following infection varies widely from asymptomatic carriage to life-threatening respiratory failure and death.

Since early in the pandemic, it was recognized that patients with severe forms of disease, e.g. requiring hospitalization or ventilation, exhibited elevated levels of inflammatory cytokines (Pedersen and Ho, 2020). This inflammatory state was associated with end-organ damage and in some cases death (Mangalmurti and Hunter, 2020; Tisoncik et al., 2012). While it remains unclear how the individual cytokines associated with this response may be involved in severe outcomes in patients, inflammation is thought to be a primary driver of later stages of this disease. In support of this hypothesis, the use of the anti-inflammatory steroid, dexamethasone, decreased mortality by 29% in COVID-19 patients who required mechanical ventilation (The RECOVERY Collaborative Group, 2020).

Aligned with these clinical observations, efforts to characterize the host response to infection and identify contributors to severe clinical outcomes have been ongoing since the pandemic began. Proinflammatory mediators such as the cytokines interleukin-6 (IL-6) and TNF-α have been associated with severe disease. Cytokine-targeted therapies have been proposed and in some cases are in clinical trials. For example, the recently completed Adaptive COVID-19 Treatment Trial 2 (ACTT-2) showed a faster time to recovery with the Janus kinase inhibitor baracitinib plus remdesivir compared to remdesivir alone (Kalil et al., 2020), as well as ACTT-4 which is comparing baracitinib plus remdesivir vs dexamethasone plus remdesivir (ClinicalTrials.gov Identifier: NCT04640168).

Descriptive studies of the immune response to SARS-CoV-2 have shown it to be highly heterogeneous (Del Valle et al., 2020; Liao et al., 2020; Nienhold et al., 2020), including the observations that CD4^+^ T cells from COVID-19 patients secreted the Th1 cytokine IFN-γ, the Th17 cytokines IL-17A and IL-17F, and the Th2 cytokine IL-4 (Su et al., 2020; Weiskopf et al., 2020). This level of diversity and variability make it especially challenging to find specific drivers of disease and options for therapies. Consequently, understanding the mechanisms by which distinct immune responses contribute to COVID-19 severity will be crucial to designing personalized or targeted interventions, and ultimately to improve upon the current steroid-based treatments.

In this study, we characterized the immune response of patients with COVID-19 and identified the type 2 cytokine, IL-13 as associated with severe outcomes. Using a mouse model of COVID-19, we discovered that IL-13 promotes severe disease, and that this response is likely to be at least partially mediated by the deposition of hyaluronan in the lungs.

## Results

### IL-13 is associated with severe COVID-19 in two patient cohorts

We analyzed plasma cytokines in 178 patients with COVID-19 at the University of Virginia Hospital, 26 of whom received their care as outpatients and 152 as inpatients (**Table S1**). Cytokines were measured in the blood sample taken closest to the first positive COVID-19 RT-qPCR test (**Figure S1**). To understand the potential interrelationships between the different cytokines measured in our cohort, we generated a heatmap for patients, grouped by hospitalization and ventilation status and cytokine expression (**Fig. 1A**). While ventilated patients appeared to have elevated levels (dark purple) of many the measured cytokines, there was a high degree of heterogeneity among the patients. Cytokines were arranged by principal component 1 (PC1) from **Fig 1B**.

**Figure 1.**
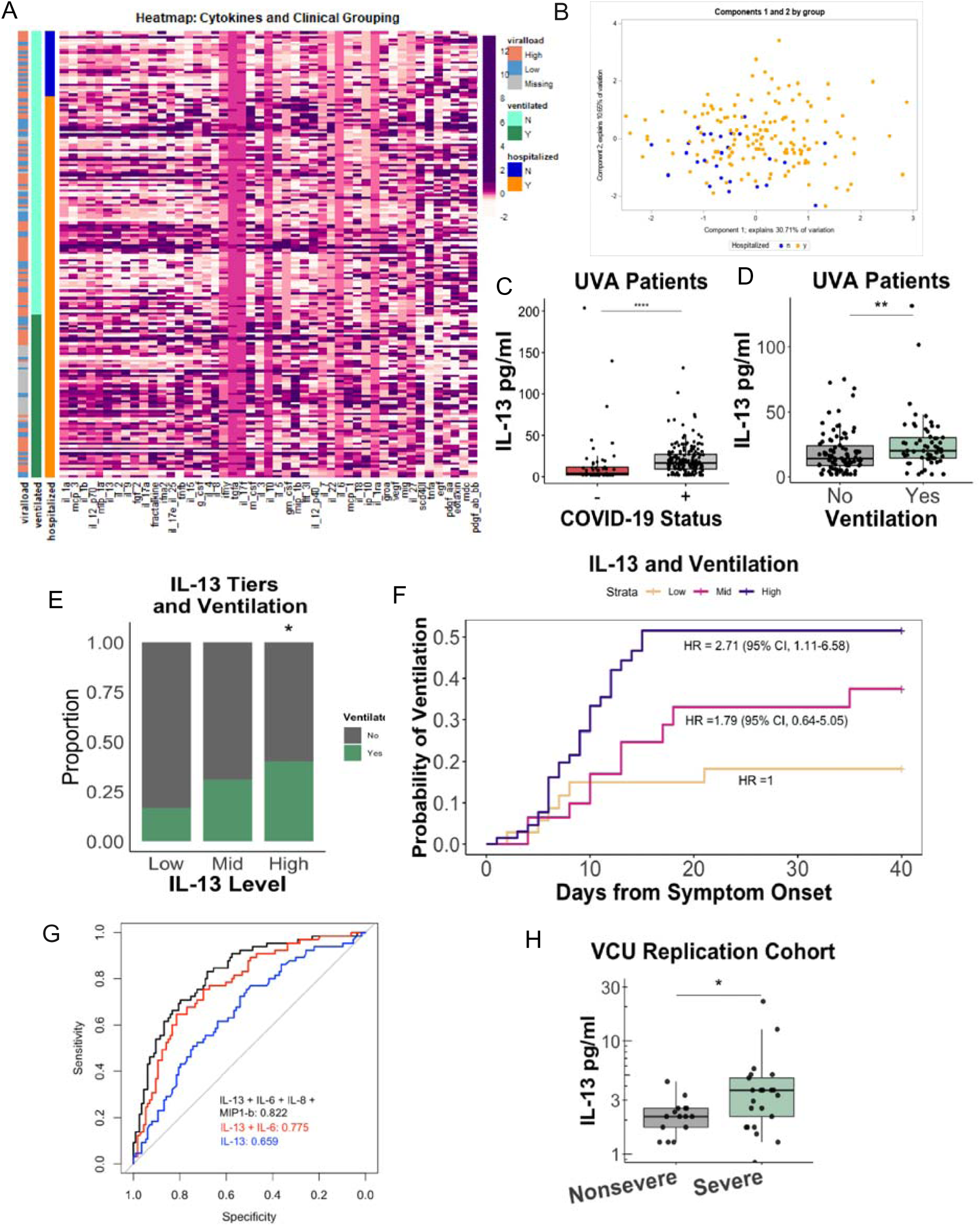
Type 2 immune response in patients with severe COVID-19 disease. (A-E) Cytokines were measured in plasma from 26 outpatients and 152 inpatients with COVID-19 infection at the University of Virginia Hospital using a 48-plex cytokine array. A) Heatmap of plasma cytokines, supplemental oxygen requirement and nasopharyngeal viral load, with rows ordered by patient status (outpatient (OP) vs inpatient (IP)) and columns by cytokine principal component 1 which included IL-13 (Table S2). B) Scatterplot comparing principal component 1 and 2 from Principal Component Analysis (PCA) of the plasma cytokines (orange inpatients and blue outpatients). C) Plasma IL-13 levels in COVID-19 patients who were or were not diagnosed with COVID-19 or D) did or did not require mechanical ventilation (Wilcox test). E) Proportion of COVID-19 patients requiring mechanical ventilation stratified by IL-13 plasma cytokine levels (Chi-square analysis). F) Kaplan-Meier survival analysis of the relationship between IL-13 level and mechanical ventilation. Comparison made to lowest IL-13 quantile (Logrank; Cox proportional hazards test adjusted for age, sex, and comorbidities). G) ROC curve with AUC plotted from: IL-13 alone (blue), IL-13 and IL-6 (red), or IL-13, IL-6, IL-8, and MIP-1b (black). G) IL-13 levels in 19 non-severe and 26 severe (requiring supplemental oxygen) COVID-19 patients from Virginia Commonwealth University Hospital (Wilcox test). * =p<0.05; ** =p< 0.005.

A principal component analysis (PCA) of the patients based on the cytokines measured was performed and the scatterplot of components 1 and 2 showed separation, albeit with overlap, of outpatients with less severe disease (**Fig 1B, blue dots),** from inpatients (**Fig. 1B, yellow dots**). IL-13 was in the top 6 of the 47 cytokines/growth factors in component 1 of the PCA (**Table S2**). This is of importance because the components estimated via PCA are able to retain information, separating patients by disease severity (inpatient vs outpatient). IL-13’s position as a high-ranking contributor to component 1, suggested its overall importance in the disease process. Additionally, network analysis of all of the cytokines measured showed the close relationship between IL-13 and other type 2 cytokines identified in component 1 (**Fig. S2**).

IL-13 is implicated in numerous processes, including (i) recruitment of eosinophils and M2 macrophages to the lung, (ii) induction of mucus secretion into the airways and goblet cell metaplasia, (iii) proliferation of smooth muscle cells, and (iv) fibrosis via fibroblast activation and subsequent collagen deposition (Chiaramonte, 2001; Marone et al., 2019). Therefore, IL-13, as an integral orchestrator of pathogenic responses in the lung, was of particular interest to us. Plasma levels of IL-13 were significantly higher in COVID-19 positive patients compared to uninfected patients (**Fig 1C**), consistent with previous reports (Huang et al., 2020; Petrey et al., 2020; Yang et al., 2020). Coordinately, we found plasma IL-13 levels to be significantly elevated in patients who required mechanical ventilation (**Fig. 1D**). Additionally, we stratified patients into three IL-13-expression level groups, and found the high-expressing tier was associated with ventilation as shown by a Chi-Square analysis as well as increased probability of ventilation (HR 2.72; 95% CI, 1.12-6.61) (**Fig. 1E, F**). To confirm that this association with IL-13 and disease severity was not because of the possibility that patients who required ventilation had elevated cytokines due to a longer duration of illness by the time their sample was drawn, we performed a linear regression between IL-13 and days from symptom onset (**Fig S2B**), and saw no significant correlation. To assess the ability of IL-13, alone or in combination with other cytokines, to be able to predict ventilation outcomes in patients, we utilized a receiver operating characteristic (ROC) analysis. We found that IL-13 alone performed modestly (area under the ROC curve [AUC] = 0.659). Inclusion of the cytokine IL-6 increased the predictive capability (AUC = 0.775), and additional inclusion of IL-8 and MIP-*β* further improved the model (AUC = 0.822) (**Fig 1G**). To validate results from the University of Virginia Hospital, cytokines from an additional 48 inpatients with COVID-19 from Virginia Commonwealth University Medical Center were analyzed (**Fig. S3; Table S3**). IL-13 levels measured in plasma were found to be elevated in the hospitalized patients who received oxygen via high flow nasal canula or mechanical ventilation compared to inpatients who did not (**Fig. 1H**).

Consistent with our study, Lucas *et al*. 2020 found that IL-13 increased from day 5 to 20 of illness in severe COVID-19 patients requiring ICU and/or mechanical ventilation. Together, these data highlight that IL-13 may be an important component of host responses to SARS-CoV-2 infection and could be driving severe disease.

### Type 2 immunity is induced in k18-hACE2 mice

To test the contribution of IL-13 to respiratory failure in COVID-19, we utilized a K18-hACE2 transgenic mouse model of COVID-19 (Moreau et al., 2020; Rathnasinghe et al., 2020; Winkler et al., 2020). In this model mice progress to severe disease starting at day five post-infection (pi) with SARS-CoV-2.

We characterized the impact of SARS-CoV-2 infection in the mouse lung. Reactome analysis of differentially regulated genes from whole tissue RNAseq revealed a significant enrichment of genes involved in IL-4 and IL-13 signaling in infected lung (FDR= 0.03) on day five pi (**Table S4**). This included the receptors through which IL-13 signals, *Il4ra* and *Il13ra1*, although *Il4* and *Il13* transcripts were not differentially expressed by day five of infection. Importantly, the receptors through which IL-13 signals, *Il4ra* and *Il13ra1*, were upregulated, indicating that potential for increased signaling even in the absence of detectable increases in cytokine expression. Additional type 2 immune effectors known to be regulated by IL-4 and IL-13 including *Chil3*, *Retnla*, *Ccl11* and *Arg1* were impacted by SARS-CoV-2 infection (**Fig. 2A**).

**Figure 2.**
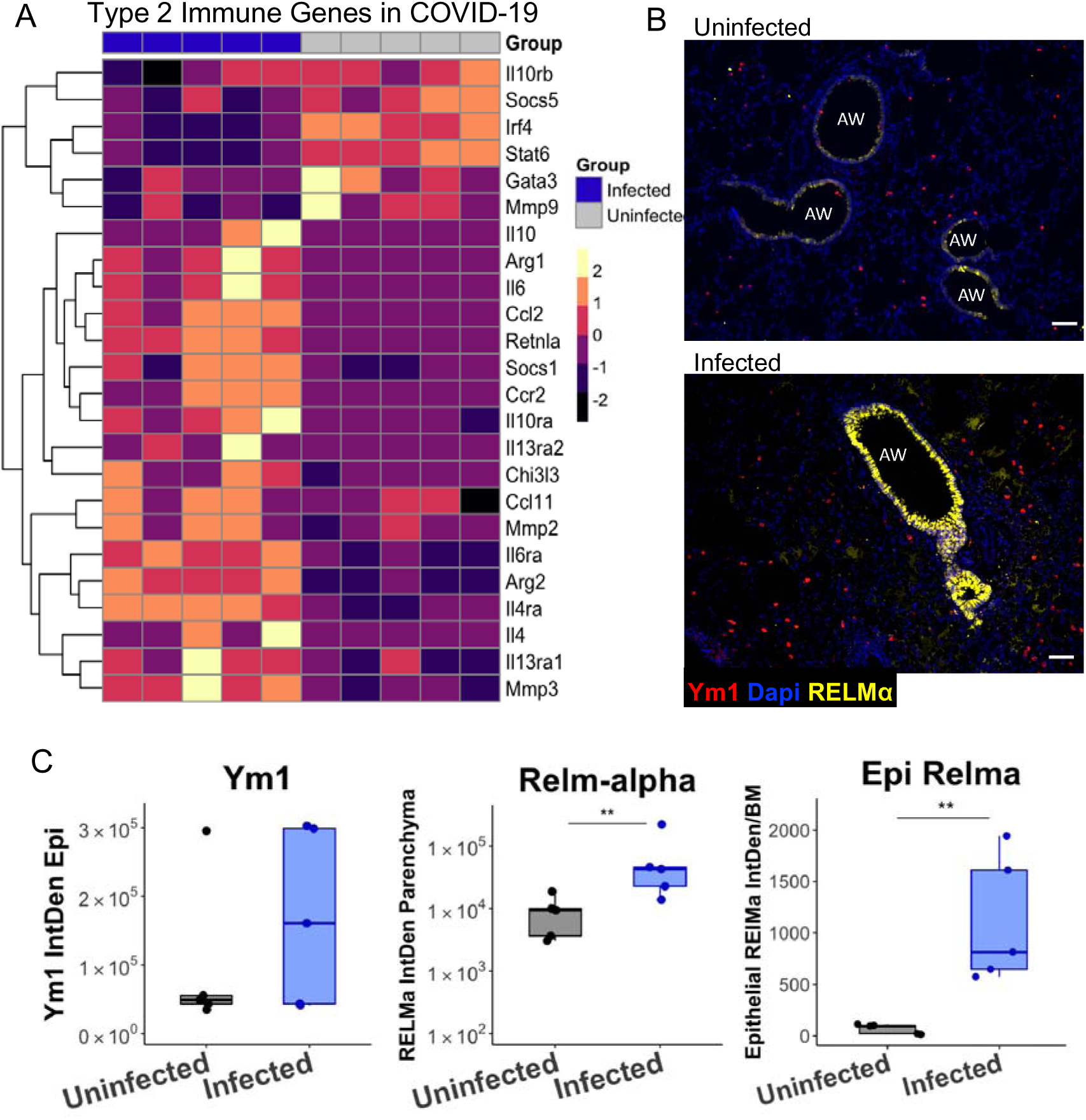
Type 2 immune response in lungs of mice following infection with SARS-CoV-2. 10-week-old male mice (Tg K18-hACE2 2Prlmn) were infected with 5×10^3^ PFU of SARS-CoV-2 and lung tissue examined on day five post-infection by RNA-seq and immunohistochemistry (IHC). A) Type 2 gene expression in the lungs of infected vs uninfected mice (heat map of normalized values of manually curated list of type 2 immune pathway genes). B) Immunohistochemistry of the type 2 immunity proteins RELMa (RELMa) and Ym1 in the lungs of infected and uninfected mice (AW, airway). C) Quantification of IHC scoring for RELMa and Ym1 (mixed effect model). *=p<0.05; **=p<0.005

In support of enhanced type 2 associated genes, protein expression of Ym1 (*Chil3*) and RELMα (*Retnla*) was increased in the lungs following infection (**Fig. 2B,C**), as measured by immunohistochemistry (IHC). Together, this highlighted that type 2 immunity was induced in the lungs due to infection. It is important to note that IL-4 and IL-13 share a receptor subunit and induce common pathways, so it is difficult to delineate their respective contributions to the upregulation of type 2 effector genes (McCormick and Heller, 2015). However, because we observed an association of IL-13 but not IL-4 (**Fig S1**) with severe disease in patients, we hypothesized that IL-13 signaling in the lung following infection was contributing to worse outcomes.

### IL-13 neutralization in mice reduces disease severity

To directly test whether IL-13 was deleterious following SARS-CoV-2 infection, we administered intraperitoneal (i.p.) injections of anti-IL-13 or an isotype matched control IgG on days 0, 2 and 4 pi. Infected mice receiving anti-IL-13 had significantly reduced symptoms as measured by clinical scores (Moreau et al., 2020) (**Fig. 3A**), weight loss (**Fig. 3B**) and mortality (**Fig. 3C**), demonstrating a pathogenic role for this cytokine during disease. Importantly, anti-IL-13 did not alter viral load in the lungs on day five (**Fig. S3A**), suggesting disease amelioration was not due to reduced infectious burden but likely due to events downstream of IL-13 signaling.

**Figure 3.**
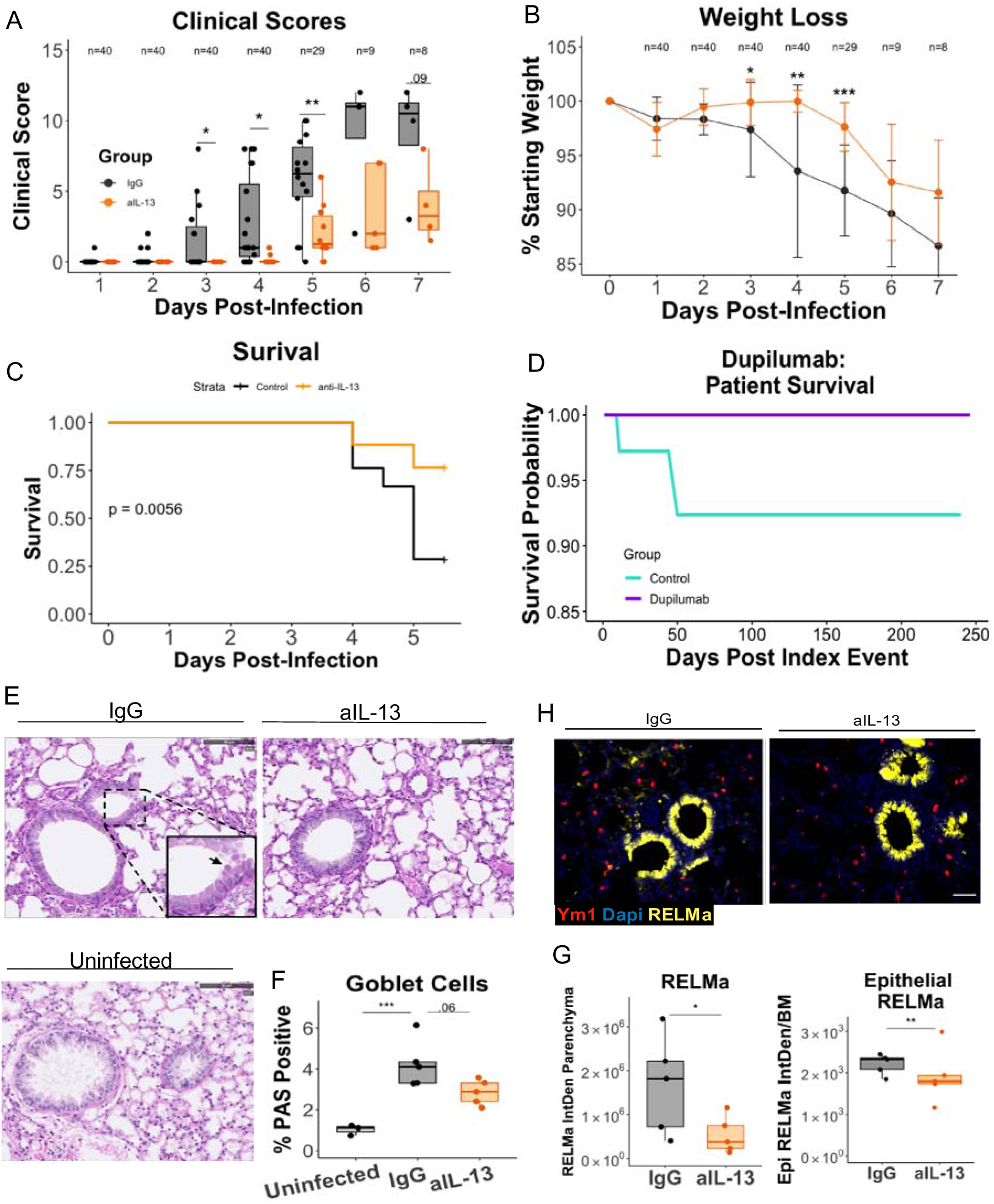
IL-13 neutralization protects from severe COVID-19 in K18-hACE2 mice. Mice were infected on day 0 with 5×10^3^ PFU of SARS-CoV-2 and administered 150 µg of anti-IL-13 or an IgG isotype control antibody intraperitoneally on days 0, 2, and 4. A) Clinical scores of illness severity on days 1-7 pi. Clinical scoring was measured by weight loss (0-5), posture and appearance of fur (piloerection) (0–2), activity (0–3) and eye closure (0–2). B) Weight loss on days 1-7 pi. C) Kaplan-Meier survival analysis in mice. D) Kaplan-Meier curve generated from data obtained from TriNetX: 1:1 matching based on 81 patients who had been prescribed Dupilumab independently of their COVID-19 diagnosis. E) Histopathology of lung tissue comparing infected mice with or without IL-13 neutralization to uninfected mice (PAS stain; inset arrow indicates PAS (+) goblet cell. F) Quantification of scoring for goblet cell metaplasia in the IgG isotype control and anti-IL-13 treatment groups. G) Immunohistochemistry of lung tissue stained for RELM-a (yellow) in parenchyma or airway, and DAPI (blue).H) Quantification of intensity of staining for RELM following IL-13 neutralization (log transformed, mixed effect model). (N = 5 mice/group; A & B combined from three independently conducted experiments). *=p<0.05; **=p<0.005.

Dupilumab is a monoclonal antibody that blocks IL-13 and IL-4 signaling. It is directed against the IL-4Rα subunit that is shared with the IL-13 receptor (Le Floc’h et al., 2020). Based on our findings, we considered the possibility that patients prescribed Dupilumab for asthma, atopic dermatitis or allergic sinusitis may be protected from severe COVID-19. To test this hypothesis, we conducted a retrospective analysis of a large international COVID-19 cohort comprised of 350,004 cases; 81 of which had been prescribed Dupilumab prior to, and independently of their COVID-19 diagnosis. We generated a sub cohort using 1:1 propensity score matching as well as an additional sub cohort for patients with diagnosed asthma, atopic dermatitis or rhinosinusitis, for which Dupilumab is prescribed. Importantly, Dupilumab use was associated with a lower risk of ventilation and death in COVID-19 (**Fig 3D; Table S5-S7**). To assess whether Dupilumab was associated with clinical proxies of inflammation, we also examined levels of C-reactive protein (CRP), an acute phase protein that increases during inflammation and correlates with poor outcomes in COVID-19 (Liu et al., 2020). CRP levels were reduced in Dupilumab-prescribed patients with COVID-19 (**Table S8**), suggesting that blocking type 2 immunity may lower over-all disease pathology and increase survival rates. As a validation cohort we also examined Dupilumab and COVID-19 in the National COVID Cohort Collaborative (N3C). The 31 patients who contracted COVID-19 while on Dupilumab also had a lower rate of hospitalization, ventilation, and death compared to matched controls (**Table S9**).

To further understand the potential mechanism by which IL-13 exacerbated disease we used the mouse model to ask whether the reduction in disease severity following IL-13 neutralization corresponded with changes in the lung tissue. We therefore assessed histological parameters of pathology by H&E staining. We have previously reported that infection with SARS-CoV-2 in this model results in lung damage (Moreau et al., 2020), however, IL-13 blockade resulted in little change in lung injury (**Fig. S3B,C**). In contrast, goblet cell metaplasia was subtly, albeit significantly, induced following infection, and this increase was reduced by neutralization of IL-13 (**Fig. 3E,F**). Lastly, major changes in collagen deposition were not observed with Masson’s trichrome stain due to IL-13 blockade (**Fig. S3D,E**) over the time period assessed. Our results provide evidence that IL-13 signaling is active in COVID-19 but less dramatic histologically than in other models of type 2 immunity (Borthwick et al., 2016; Lee et al., 2011).

To further investigate how IL-13 neutralization protected from COVID-19, we assessed expression of the type 2 associated proteins Ym1 and RELMα. Fluorescence microscopy of the lung revealed a significant decrease in RELMα following neutralization of IL-13, which was evident in both the parenchyma and within the epithelial cells (**Fig. 3G,H**). However, no change in Ym1 following IL-13 neutralization was detected (**Fig. S3F)**. In addition, no overt changes in cytokine levels or cell composition in the bronchoalveolar lavage fluid were observed (**Fig. S3G,H**). We concluded that the mechanism by which IL-13 was promoting more severe COVID-19 was not necessarily through the type 2 pathways typically observed in the lung.

### Hyaluronan is associated with severe COVID-19

Given the above, we took a more unbiased approach to evaluate the impact of IL-13 during COVID-19. RNA-seq analysis was performed on whole lung tissue from IL-13-neutralized and control-treated mice at day five pi to evaluate transcriptional responses downstream of IL-13. Intriguingly, the transcriptomics analysis identified the enzyme hyaluronan synthase 1, *Has1,* which was upregulated during infection, as the most downregulated gene following IL-13 neutralization (**Table 1**, **Fig. 4A**). In addition to *Has1,* other genes associated with the hyaluronan (HA) pathway, *Cd44* (**Fig. 4A**) and *Has2* (**Fig. S4A**) were also downregulated following IL-13 neutralization. Additionally, hyaluronidases, which can break down high molecular weight (HMW) hyaluronan into the potentially more proinflammatory low molecular weight (LMW), were upregulated during infection (**Fig S4B**) supporting a potentially pathogenic role for hyaluronan during COVID-19. Deposition of the polysaccharide hyaluronan was found to be significantly increased in SARS-CoV-2 infected compared to uninfected mice, specifically in the parenchyma of the lungs (**Fig 4E,F**). Following IL-13 neutralization in infected mice, hyaluronan deposition was significantly reduced in the parenchyma (**Fig 4E,F**), supporting data that demonstrated IL-13 regulated gene expression of HA synthases.

**Table 1.**
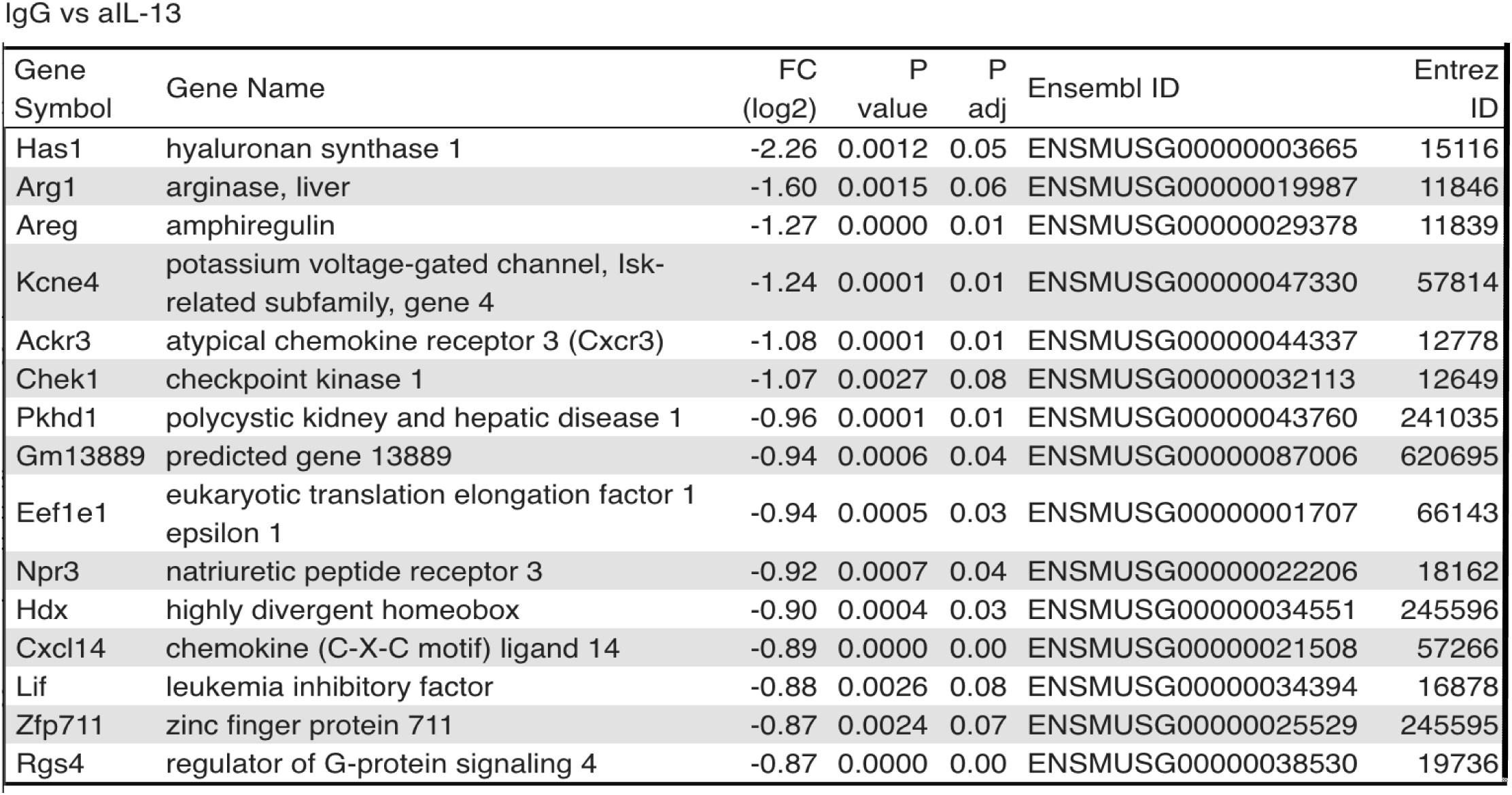
Candidate IL-13-regulated pathogenic effectors: top 15 protein coding genes downregulated by anti-IL-13 treatment relative to IgG isotype on day five of murine Covid-19 infection.

**Figure 4.**
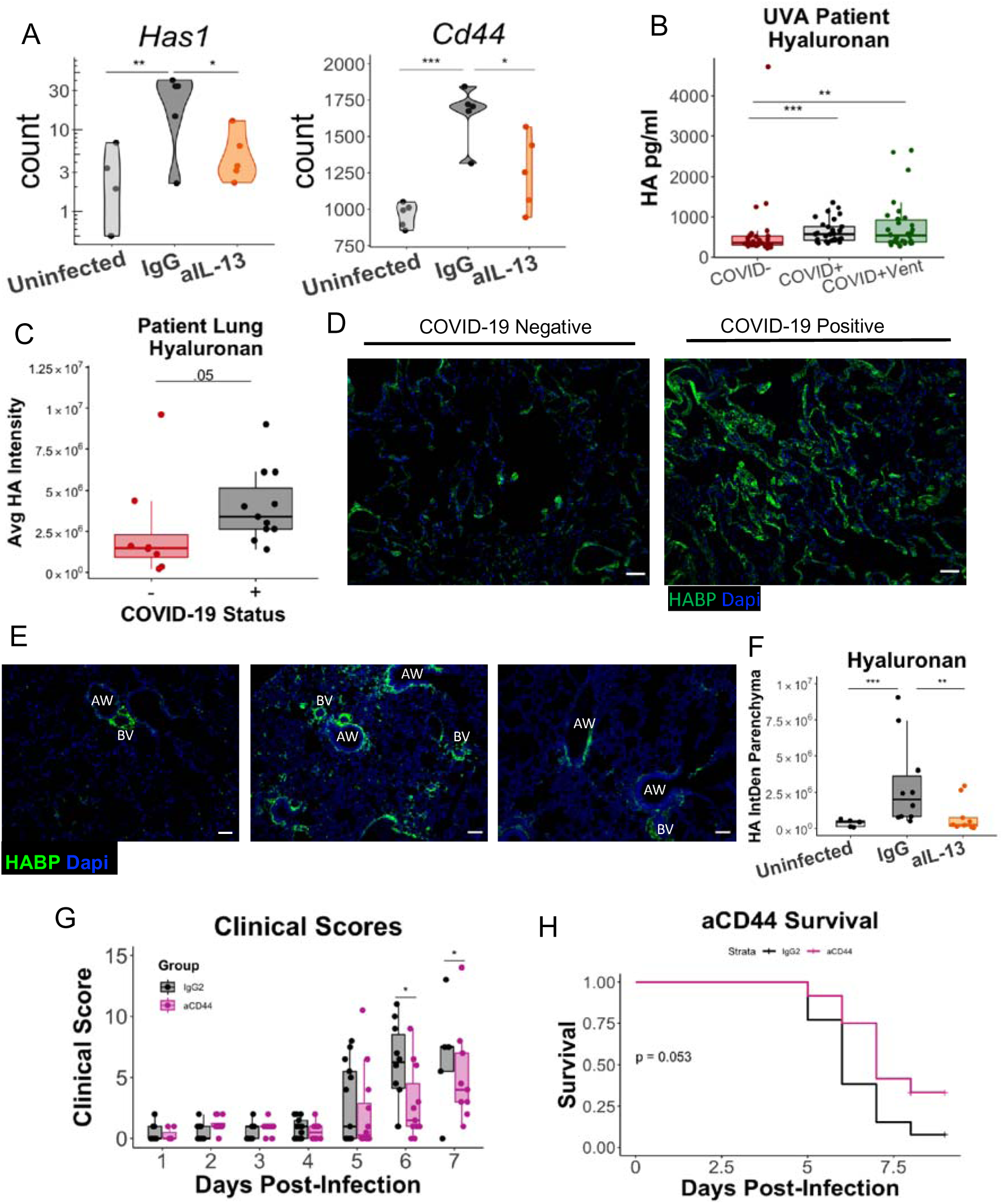
Hyaluronan and COVID-19 disease. Mice received i.p. injections of anti-IL-13 on days 0 and 2 pi, were euthanized on day 5 and lung tissue was split and placed either into trizol tissue reagent for RNA analysis or formaldehyde for paraffin embedding and immunohistochemistry. A) Gene expression in mouse lung of hyaluronan synthase (*Has1)* and the hyaluronan receptor *Cd44* of infected mice with anti-IL-13, isotype control antibody and uninfected controls. B) Hyaluronan was measured in the plasma of COVID-19-negative controls and in patients with COVID-19 that did or did not require supplemental oxygen. Postmortem lung samples were obtained from fatal COVID-19 cases and control tissue from COVID-19 negative deaths. C) Quantification of hyaluronan deposition in fatal COVID-19 disease (N=11) and controls (N=8) (log transformed, mixed effect model) using hyaluronan-binding protein (HABP). D) Representative images of staining for hyaluronan (with HABP). E) Staining of hyaluronan in mouse lung (with HABP) and F) quantification of hyaluronan deposition in tissue following infection and neutralization of IL-13 (mixed effect model; combined 2 experiments). Mice were administered anti-CD44 or IgG2 isotype control on days 1, 2, 3 and 4 pi. G) Clinical scores and H) Kaplan-Meier survival curve for mice; combined two, independent experiments. *=p<0.05; **=p<0.005; ***=p<0.0005.

Our finding in the mouse model that IL-13 regulates the HA pathway in COVID-19 was noteworthy as there is evidence to support a pathological role for HA in humans with lung disease (Bell et al., 2019) including COVID-19 (Ding et al., 2020; Hellman et al., 2020; Nagy et al., 2019; Ruppert et al., 2014; Rayahin et al., 2015). We observed that patients infected with SARS-CoV-2 had higher plasma levels of hyaluronan (**Fig 4B**). Additionally, hyaluronan was elevated in the postmortem lung tissue from patients who died of severe COVID-19 (**Fig 4C,D; Fig S4H**), supporting prior observations (Hellman et al., 2020). It is important to note, however, that hyaluronan can be elevated due to an array of factors, including occupation, respiratory diseases, or other infections unrelated to SARS-CoV-2 (Lauer et al., 2015), which likely accounts for the variation in COVID-19 negative patients.

In a murine influenza model, administering hyaluronidase to break down pathogenic HA resulted in ameliorated disease (Bell et al., 2019). We followed a similar approach to test the impact of hyaluronan during COVID-19, by administering hyaluronidase intranasally on day five pi. This resulted in modest, albeit not statistically significant, protection from weight loss and clinical scores (**Fig. S4D-F**). This suggested that IL-13-dependent accumulation of hyaluronan may not act alone to drive severe disease. Importantly, however, ovine hyaluronidase used here also reduces chondroitin sulfate (Honda et al., 2012), which may have unpredictable effects. We therefore pursued this observation by testing if blockade of the hyaluronan receptor CD44, which was also downregulated by IL-13 neutralization, would be protective. This receptor is present on multiple cell types, including inflammatory cells, that may utilize CD44-HA interactions for migration, activation, proliferation, or other functions that could contribute to pathogenic responses (Misra et al., 2015; Heldin et al., 2020). Blockade of CD44 from days one through four post-infection resulted in improved clinical scores and survival (**Fig. 4G,H, Fig. S5G**). Together, these indicated that hyaluronan and its interacting components are downstream of IL-13 and contribute to disease outcome.

## Discussion

Here, we have shown that the type 2 cytokine, IL-13, is associated with severe COVID-19. The IL-13 blocking drug, Dupilumab, in turn is associated with better outcomes in COVID-19 patients and additionally, neutralization of IL-13 in mice infected with SARS-CoV-2 protects from death, in part by blocking hyaluronan synthesis and excessive deposition. Overall, this work opens a new avenue in the study of COVID-19 by demonstrating a causal role for type 2 immune responses and downstream hyaluronan accumulation in respiratory failure, and offers potential avenues for immunotherapy of this disease. Considering the extreme heterogeneity in immune responses to COVID-19 it is unlikely that IL-13 blockade will work in all patients. Nonetheless, understanding the underlying mechanism and identifying those most likely to benefit from such a treatment would be a major advance.

The mechanism through which IL-13 promoted severe disease was challenging to identify, as there was only a modest impact on the downstream effectors of IL-13 that are commonly seen during allergic or asthmatic inflammation. Although there were decreases in type 2 associated responses, e.g. goblet cells and RELM*α*, following IL-13 blockade, the biological significance of their contribution was likely minimal given the low magnitude of their induction compared to other models of type 2 immunity in the lung. The identification of *Has1* as the most downregulated gene following IL-13 neutralization in infected mouse lungs, along with downregulation of *Has2* and *Cd44,* two other genes involved in the HA pathway, enabled the discovery of a novel route by which IL-13 impacts pathology via upregulation of hyaluronan synthesis. We showed that IL-13 neutralization not only decreased *Has1* gene expression but lowered hyaluronan deposition in the lung. Downstream of hyaluronan production, neutralization of the hyaluronan receptor improved survival in infected mice. While our work stops short of a full mechanistic understanding of the function of hyaluronan in COVID-19, it is interesting to speculate that this polysaccharide may contribute to inflammation in the lung by providing a matrix for inflammatory cells to migrate over and adhere to, as well as via signaling through its receptor CD44. Additionally, excessive build-up of hyaluronan, which binds a large amount of water, could contribute to severely impaired oxygen uptake, which is a significant component of disease in hospitalized patients.

Because increases in hyaluronan have been observed in patients with COVID-19, this study provides a potential mechanistic link between the association of IL-13 with severe disease (Lucas et al., 2020) and increased hyaluronan seen in other studies (Ding et al., 2020; Hellman et al., 2020). Understanding the relationship between IL-13 and HA may be widely relevant to respiratory diseases beyond COVID-19.

Overall, this work in humans and mice implicates IL-13 as an important driver of severe outcomes during COVID-19, in part through hyaluronan-CD44 engagement in the lungs. Understanding the pathogenic role of IL-13 in the murine model, combined with the results correlating Dupilumab use with better patient outcomes, emphasizes the potential impact this work may have on improving patients’ lives.

## Materials and Methods

### Study Design

Discarded human plasma samples from COVID-19 positive and negative patients at the University of Virginia Medical Center were collected for cytokine and growth factor analyses. The collection of biological specimens and de-identified patient information was approved by the University of Virginia Institutional Review Board (IRB-HSR #22231 and 200110). In mice, neutralizing antibodies or isotype controls were used to assess the role of IL-13 during COVID-19 All mouse work was approved by the University of Virginia Institutional Animal Care and Use Committee, and all procedures were performed in the University certified animal Biosafety Level Three laboratory.

### Patients

Patients with a positive RT-qPCR test for COVID-19 at the Clinical Microbiology Laboratory at the University of Virginia Medical Center had any remnant EDTA-plasma samples within 48 hours of the time of diagnosis or hospitalization collected. EDTA-plasma from 178 patients diagnosed from March to September 2020 were analyzed in this study. Blood was centrifuged at 1300 x *g* for 10 minutes, then plasma was aliquoted and stored at −80°C until testing. Demographics (age, gender, race), comorbidities, hospitalization status, lab results, and other clinical information were obtained from the electronic medical record (EMR) (**Table S1**). Severity of COVID-19 illness was assessed through review of the electronic medical record in two ways: first by inpatient admission vs outpatient care, and second by the use of supplemental oxygen (none vs any supplemental oxygen, and supplemental oxygen delineated as low flow nasal canula vs mechanical ventilation or high flow oxygen). In addition, supplemental oxygen was scored as occurring at the time of the blood draw or at a future time. Days from symptom onset was scored as per Lucas *et al* (Lucas et al., 2020) based on the patient’s determination or by the earliest reported symptom from the patient as recorded in the electronic medical record.

Cytokine concentrations in plasma were measured using the MILLIPLEX® MAP Human Cytokine/Chemokine/Growth Factor Panel A (48 Plex) (Millipore Sigma, St. Louis) by the Flow Cytometry Facility of the University of Virginia. Cytokines detected were sCD40L, EGF, Eotaxin, FGF-2, Flt-3 ligand, Fractalkine, G-CSF, GM-CSF, GROα, IFNα2, IFNγ, IL-1α, IL-1β, IL-1ra, IL-2, IL-3, IL-4, IL-5, IL-6, IL-7, IL-8, IL-9, IL-10, IL-12 (p40), IL-12 (p70), IL-13, IL-15, IL-17A, IL-17E/IL-25, IL-17F, IL-18, IL-22, IL-27, IP-10, MCP-1, MCP-3, M-CSF, MDC (CCL22), MIG, MIP-1α, MIP-1β, PDGF-AA, PDGF-AB/BB, TGFα, TNFα, TNFβ, and VEGF A (**Fig. S1**). (RANTES was excluded). Hyaluronan was measured using the Hyaluronan Duoset ELISA (R&D, cat# DY3614-05) with plasma diluted 1:50.

As a test of validation of the results from the University of Virginia Hospital, an additional 47 patients with symptomatic COVID-19 (all inpatients) from Virginia Commonwealth University Medical Center were analyzed. Severe was defined as any patient requiring high flow nasal cannula (HFNC) oxygen delivery, intubation, or who’s disease resulted in sepsis or death. IL-13 was measured in EDTA plasma from these patients using the Bio-Plex Pro Human Cytokine 27-plex Assay (R & D Systems, Minneapolis, MN) (**Table S4**).

### Database and Inclusion Criteria for Dupilumab

Data were retrieved from the COVID-19 Research Network provided by TriNetX, comprising 400 million patients from 130 health care organizations in 30 countries (database access on 12/05/2020). COVID-19 patients were identified via the ICD-10 code U07.1 or the presence of a SARS-CoV-2-related RNA diagnosis within the last eleven months. Propensity scores matched cohorts 1:1 using a nearest neighbor greedy matching algorithm with a caliper of 0.25 times the standard deviation. Outcomes were defined as ventilation assist and death. Measures of association including risk differences with their respective 95% CI’s were calculated. In addition, Kaplan-Meier curves were generated for each analysis.

A sub cohort with indications for Dupilumab use was generated using ICD-10 codes for asthma (J45), atopic dermatitis (L20.8), and pansinusitis (J01.40 and J32.4). Drug use was identified via RxNorm codes for Dupilumab (1876376) and the lab value for C-reactive protein (9063).

Patients receiving Dupilumab are on average one year older.

### N3C Dupilumab Analysis

We utilized deidentified data in the National Cohort Collaborative Cohort (N3C) enclave, which currently contains 2 years of medical record data from 34 United States sites, to explore the association between Dupilumab use and COVID-19 outcomes. This enclave represents >2M persons (including ∼300K COVID-19 cases) and medical facts from more than ∼90M visits. Values less than 20 have been suppressed as per current N3C publication policy.

#### Cohort Definitions

1. Dupilumab+: If Patient had Dupilumab within 61 days prior to their first COVID-19 diagnosis date
2. Controls+: Patients with no record of Dupilumb within 2 months prior to their first COVID-19 diagnosis date.

#### Outcome Definitions

1. Hospitalized: Patients who became inpatient within 6 weeks post COVID-19 diagnosis date.
2. Death: If subjects death date is after their first COVID-19 diagnosis date.
3. With Ventilation: If patients were put on a ventilator within 6 weeks of any of their COVID-19 diagnosis dates (window is double sided: procedure could have been 6 weeks before or after any of their diagnosis dates).

The incidence of COVID positivity in people on dupilumab [cohort definition 1 above / (definition 1 + 2)], along with 95% confidence intervals, was calculated. Then, a case-control design was used. Dupilumab+ patients were matched to control+ patients in a 1:5 ratio, with exact matching on gender, race, ethnicity, N3C site, asthma and nearest matching on age.

Conditional logistic regression was used to compare COVID-19 severity outcomes within this matched subset of COVID+ patients. A sensitivity analyses was performed excluding asthma from the matching criteria.

### Virus and Cell Lines

SARS-Related Coronavirus 2 (SARS-CoV-2), isolate Hong Kong/VM20001061/2020 (NR-52282) was obtained from the Biodefense and Emerging Infections Research Resources Repository (BEI Resources), National Institute of Allergy and Infectious Diseases (NIAID), National Institutes of Health (NIH). Virus was propagated in Vero C1008, Clone E6 (ATCC CRL-1586) cells cultured in Dulbecco’s Modified Eagle’s Medium (DMEM, Gibco 11995040) supplemented with 10% fetal bovine serum (FBS) and grown at 37°C, 5% CO_2_. Initial viral stocks were used to infect Vero E6 cells, generating passage 1 (P1) stocks. These P1 stocks were then used to infect additional Vero E6 cells, generating passage 2 (P2) stocks, which were used for all experiments.

### Viral Propagation

Vero E6 cells grown to 90% confluency in T75 tissue culture flasks (Thermo Scientific) were infected with SARS-CoV-2 at a multiplicity of infection of 0.025 in serum-free DMEM. Vero E6 cells were incubated with virus for two hours at 37°C, 5% CO_2_, after which the virus was removed, media was replaced with DMEM supplemented with 10% FBS, and flasks were incubated at 37°C, 5% CO_2_. After two days, infected flasks showed significant cytopathic effects, with >50% of cells unattached. Cell supernatants were collected, filtered through a 0.22µm filter (Millipore, SLGP003RS), and centrifuged at 300 x g for ten minutes at 4°C. Cell supernatants were divided into cryogenic vials (Corning, 430487) as viral stocks and stored at −80°C until use.

#### Challenge

8-16 week-old male Tg(K18-*hACE2*) 2Prlmn (Jackson Laboratories) (Moreau et al., 2020) mice were challenged with 5000 plaque forming units (PFUs) of SARS-CoV-2 in 50 μ by an intranasal route under ketamine/xylazine sedation. Mice were followed daily for clinical symptoms, which included weight loss (0-5), activity (0-3), fur appearance and posture (0-2), and eye closure (0-2). Mice were given 150 µg of anti-IL-13 (clone eBio1316H; cat # 16-7135-85) or an isotype matched control IgG (clone eBRG1; # 16-4301-85) administered on day 0, 2, and 4 pi. For experiments utilizing hyaluronidase, 14 U in 70 µL of ovine testicular hyaluronidase (Vitrase; 200 USP Units/mL) or saline control were administered intranasally following isoflurane anesthetization on day five pi. For anti-CD44 experiments, 100 µg of anti-CD44 (BD Biosciences, clone IM7; cat # 553131) or IgG2 (BD Biosciences, clone A95-1; cat # 559478) were administered on day one pi, and then mice were given an additional 50 µg on days 2, 3 and 4.

#### Viral titers

The left lobe of the lung was removed, placed in a disposable tissue grinder with 1 mL of serum-free DMEM on ice, and then ground. Lung homogenates were centrifuged at 300 x *g* for 10 minutes, and then the supernatants were collected and frozen at −80° C until use. Plaque assays were as described previously (Moreau et al., 2020). Briefly, Vero E6 cells seeded in 12-well tissue culture plates were infected with lung homogenates serially diluted in serum-free DMEM. Plates were then incubated at 37° C, 5% CO_2_ for two hours to allow viral infection of the cells, before washing with sterile PBS (Gibco 10010-023) to remove virus and replacing with an overlay of DMEM, 2.5% FBS containing 1.2 % Avicel PH-101 (Sigma Aldrich). After incubation at 37°C, 5% CO_2_ for two days the overlay was removed, wells were fixed with 10% formaldehyde and stained with 0.1% crystal violet to visualize plaques and calculate viral titers as PFU/ml.

#### Histology

Tissues were fixed in formaldehyde, processed and embedded in paraffin. Sections were stained with H&E, Masson’s Trichrome and Periodic Acid-Schiff using standard protocols. Slides were scanned at 20X magnification. Histopathological scoring for lung tissue was done according to the guidelines of the American Thoracic Society (Matute-Bello et al., 2011). For antibody and HA staining, lung sections were deparaffinized and heat-mediated antigen retrieval performed using Tris-EDTA buffer (10 mM Tris Base, 1 mM EDTA, 0.05 % Tween-20 pH 8.0; incubation 20 min 95°C). Non-specific protein was blocked (2% donkey serum, 1% BSA, 0.05% Tween-20) prior to blocking endogenous avidin and biotin (Thermofisher). Lung sections were incubated with primary antibodies or HA binding protein (**Table S10**) overnight at 4°C, washed in PBS containing 0.05% Tween-20 before incubation with secondary antibodies (**Table S10**) for 1 hr at room temperature followed by mounting with DAPI containing fluoromount (Southern biotech). Images were captured with an EVOS FL imaging system (Thermofisher). Analysis of images was performed using ImageJ software (version 2.09.0-rc69/1.52p) on sections where sample identification was blinded for the investigator. For calculation of antibody positive staining, background autofluorescence was subtracted from all images based on pixel intensities of sections stained with secondary antibodies only. Analysis was limited to regions of interest (airway, vessels or parenchyma), a threshold was applied to all images to incorporate positively stained pixels, and area of positively stained pixels within the region of interest calculated. For measurements of airway positivity, stain intensity was normalized to the length of the basement membrane.

### Mouse bronchioalveolar lavage

BAL fluid was collected from each animal through cannulation of the exposed trachea, and flushing twice with .7mL of PBS. BALF samples were centrifuged at 500xG for 5 minutes, and supernatant was immediately frozen for later cytokine analyses. For flow cytometry, pelleted cells were resuspended in FACS (PBS + 5% FBS) buffer for staining and with Zombie NIR (Biolegend, 423105, SanDiego, CA), CD45, Alexa Fluor 532 (eBioscience, 58-0459-42,San Diego, CA), CD11c, PE-Cy7 (Biolegend, 117317, SanDiego, CA), CD11b, BV480 (BD Biosciences, 566117, SanJose, CA), SIGLEC F, AF700 (eBioscience, 56-1702-80, SanDiego, CA), Ly-6c, FITC (Biolegend, 128005, San Diego, CA),and Ly-6G, BV650 (Biolegend, 127641, San Diego, CA) and then fixed in IC-fixation buffer (eBioscience, 00-8222-49, San Diego, CA). Samples were run on a Cytek Aurora Borealis at the University of Virginia flowcytometry core. Neutrophils are Zombie NIR−, CD45+,CD11C−, CD11B+, and Ly-6G+; eosinophils are Zombie NIR−,CD45+, CD11C−, CD11B+, and Siglec-F+; inflammatory monocytes are Zombie NIR−, CD45+, CD11C−, CD11B+, Ly-6G−, and Ly-6C+.

Cytokine analyses were performed via Luminex Mouse 32-plex (MCYTMAG-70K-PX32, Millipore-sigma). Samples were run following manufacturers protocol after an 18-hour incubation before being run on Luminex® analyzer (MAGPIX®).

### RNAseq

RNA was extracted from murine lung tissue preserved in trizol by bead beating (tissuelyserII, Qiagen), followed by a phenol-chloroform extraction and RNA Isolation Kit (Qiagen). RNA quality was assessed using Agilent Tape Station RNA kit (Agilent). Library preparation, sequencing, quality control, and read mapping was performed by the Genome Analysis and Technology Core, RRID:SCR_018883. Briefly, ribosomal RNA was depleted using the rRNA depletion kit (NEB E6310). Following rRNA depletion cDNA libraries were prepared using the NEB ultra-directional library preparation kit 2.0 (NEB E7760) and indexed using the NEBNext Multiplex Oligos for Illumina (NEB E7335, E7500). Library size and purity were verified using the Agilent Tape Station D5000 HS kit (Agilent). Library concentration was measured with a qubit DNA HS assay (Invitrogen). Libraries concentrations were normalized and 15 libraries were multiplexed per run. Diluted libraries were sequenced on the Illumina Nextseq 500 using a 150 high output kit (400 Million reads, 2×75bp paired end, 150 cycles).

### RNAseq data analysis

RNAseq reads were first processed using Cutadapt (Martin, 2011) to trim the adapter sequences and then the quality of the reads was assessed by FastQC (https://www.bioinformatics.babraham.ac.uk/projects/fastqc/) and MultiQC (Ewels et al., 2016). After these processes the reads were aligned to the mouse Ensembl GRCh38.76 primary assembly using STAR v2.5.3a (Dobin and Gingeras, 2015) in a two-passing mode to generate a gene matrix for differential gene expression. Differentially expressed genes were determined using the DESeq2 package (Love et al., 2014) in Rstudio (*RStudio Team (2020). RStudio: Integrated Development for R. RStudio, PBC, Boston, MA URL* http://www.rstudio.com/.) Enrichment analysis was applied to the total gene counts using the reactome GSA package (Griss et al., 2020) in Rstudio. Enrichment analysis was applied to the total gene counts using the CAMERA algorithm (Wu and Smyth, 2012).

### Statistical methods

For the clinical study, a total of 47 cytokines, chemokines and growth factors were measured in 178 COVID-19 positive patients with blood samples. For cytokines of interest, levels at the time of COVID-19 diagnosis or admission were compared between patients with different severity of illness using a Mann-Whitney U test. For hierarchical clustering, the pheatmap function in the pheatmap library was used (R-project.org). Cytokines were scaled by row (patients), and the clustering was calculated using the complete linkage method. Statistical analyses were performed using GraphPad Prism and R.

Since these cytokine measurements were highly correlated, principal component analysis (PCA) was performed to identify distinct features among correlated cytokines, and thus reduce the dimensionality. Due to their skewed distributions and variable scales, the cytokines were log-transformed first, and then standardized with mean of zero and standard deviation of one. The first five principal components captured 62% of total variation in the cytokines. For each principal component, those cytokines with loading score of 0.5 or above were retained, showing the strength of their influences within the component. Additionally, the network analysis using the qgraph package in R was performed to characterize the complex structural relationships among cytokine measurements and optimized with graphical LASSO. The nodes represented individual cytokines, and edges represented their correlations in that highly correlated cytokines were connected closer with thick edges (https://cran.rproject.org/web/packages/qgraph/qgraph.pdf).

Survival Analysis was performed for patients from the time to ventilation from symptom onset. Those who were not ventilated were censored at 40 days. Patients were classified into 4 quartiles based on the cumulative distribution of IL-13 levels, and the probabilities of ventilation were estimated by the Kaplan-Meier method. Since the two upper quartiles had statistically identical results in the preliminary analysis, they were combined in the final log-rank test and Cox regression done for their relative performance over the lower quartiles, and were corrected for sex, age, and cumulative number of comorbidities (including: diabetes, cancer, stroke, and heart, liver, kidney, or lung diseases).

ROC curve was generated using pROC library in RStudio. IL-6, IL-8, and MIP-1b were selected from the top variables identified by conditional random forest analysis (data not shown). All models were additionally adjusted for age and sex.

For the Dupilumab study, TriNetX analytics tools were used to obtain baseline characteristics, balance cohorts with propensity score matching and analyze outcomes of interest in the final cohorts. Baseline characteristics, including demographics, diagnoses, procedures, and medication were obtained. Propensity score matching was used to balance cohorts. Propensity scores matched cohorts 1:1 using a nearest neighbor greedy matching algorithm with a caliper of 0.25 times the standard deviation. Outcomes were defined as ventilation assist and death. Measures of association including risk differences with their respective 95% CI’s were calculated. In addition, Kaplan-Meier curves was generated.

For clinical scores, weight loss, and scored histological sections of mouse studies, a two-tailed Student’s t test was used to determined statistical significance. For IHC sections with multiple quantified images per mouse or human tissue, response differences between groups (e.g., infected vs. uninfected, or IgG vs aIL-13) were evaluated in the mixed-effects model to account for within-individual correlation, and distributions were log transformed where appropriate. P value < 0.05 was considered significant.

### Online supplemental material

Fig S1 shows individual cytokine plots from UVA and VCU patients, comparing between disease status. **Fig S2** shows network analysis of all cytokines measured in UVA patients, and IL-13 linear regression with time from symptom onset to blood draw. **Fig S3** shows absence of impact on lung viral burden, as well as absence of changes in lung damage by H&E staining or collagen deposition by Mason’s trichrome staining, following IL-13 neutralization. This figure also shows heatmap of cytokine expression and immune cell composition in the BALF of infected mice after IL-13 neutralization. **Fig S4** shows additional genes altered in the lungs following IL-13 blockade in mice, as well as epithelial hyaluronan quantification from Fig 4. Additionally, this figure shows disease outcomes following hyaluronidase administration, weight loss from CD44 blockade, and representative images from each patient autopsy sample from Fig 4C&D). **Fig S5** shows representative images from each mouse lung section from Fig 3G&H, and Fig 4E&F. **Table S1** shows patient information from our UVA cohort. **Table S2** shows PC1 from PCA analysis in Fig 1. **Table S3** shows VCU patient information. **Table S4** shows results from Reactome analysis of RNAseq data from infected mice highlighting pathways enriched in infected compared to uninfected mice. This table shows the significant enrichment of the Interleukin-4 and Interleukin-13 signaling pathway. **Tables S5-S7** show results from survival analysis of COVID-19 patients with or without Dupilumab use from the TriNetX database. The results from the full cohort (**Table S5**), a subcohort of 1:1 propensity score matching (**Table S6**), and a subcohort based on comorbidity matching (**Table S7**) show prior Dupilumab use was associated with lower mortality rates. **Table S8** shows CRP levels between different stratified groupings of COVID-19 patients, showing Dupilumab use was associated with lower CRP levels. **Table S9** shows outcomes following prior Dupilumab use from COVID-19 patients within the N3C database. **Table S10** shows antibody reagents and staining concentrations.

## Data Availability

The data will be available online when the peer-reviewed manuscript is published.

## Acknowledgments

We wish to thank the families who consented for autopsy and the COVID-19 patients who provided plasma samples. We thank Mike Solga, University of Virginia Flow Cytometry Core, for cytokine measurements, Ron Grider for electronic medical record data extraction, Panwichit Tongvichit for plasma sample collection, Patcharin Pramoonjago for Biomedical Tissue Repository support, Jennifer White for IRB protocol preparation, Dr. Sanford Feldman for comparative medicine support, Dr. Desiree Marshall and Stella Pearson for the support of the pathology specimens, Indika Mallawaarachchi and Arti Patel for N3C analyses, and Nicholas R. Natale and William A. Petri for technical support.

## Funding

This work was supported by grants to WP from the Manning Family Foundation, Ivy Foundation, Henske Family and NIH R01 AI124214. CM was supported by R01 AI148518, SB by NIH R01 AI146257, JS by NIH UL1TR003015/ KL2TR003016, BB by the Department of Laboratory Medicine and Pathology, University of Washington, JA by the Wellcome Trust 106898/A/15/Z, Wellcome Trust 203128/Z/16/Z and MRC-UK MR/K01207X/2, TES by the Asthma UK & Medical Research Foundation MRFAUK-2015-302, SP, RP by German Research Foundation (DFG): PR1562/1-1, TRR295, KFO339, and JL and SR by NCATS U24 TR002306 and UVA UL1 TR003015. Portions of this work were also supported by the UVA School of Data Science and by NSF CAREER award MCB-1350957 (C. Mura).

## Author contributions

The following authors contributed in Conceptualization (AND, TS, BM, AJD, WP); Methodology (SB, MA, GM, BM); Validation (JD, GB, MS); Formal analysis (TS, CM, SP, RP, BB, JS, JM, GL), Resources (AND, MP, AM, BB); Data Curation (MY, RC); Writing - Original Draft Writing (AND); Review & Editing (all authors); Visualization (AND); Supervision (WP, JA); Funding acquisition (WP, JA, RP, SP, JS, BB); Project administration (WP).

## Competing interests

William A. Petri, Jr. receives research funding from Regeneron, Inc. which is the maker of Dupilumab. The other authors declare no competing interests.

## Data and Materials Available

We plan to submit all corresponding data and code to Dryad or NPG hub upon the acceptance of our submission.

## Notes

### Author Declarations

The collection of biological specimens and de-identified patient information was approved by the University of Virginia Institutional Review Board (IRB-HSR #22231 and 200110). All procedures performed in this study were approved by the Virginia Commonwealth University Institutional Review Board and in accordance with the 1964 Helsinki Declaration and its later amendments or comparable ethical standards. Informed consent was obtained from all participants or by their legally authorized representatives if they were unable to give consent. IRB Protocol #HM20019182

### Summary of Updates

Patient data (figure 1) was updated with new samples and analyses techniques. Additionally, figures 2-4 were added based on animal studies and supporting human data. Supplemental data was added to support these new additions.

## References

1 Babraham Bioinformatics - FastQC A Quality Control tool for High Throughput Sequence Data.

2 Bell, T.J., O.J. Brand, D.J. Morgan, S. Salek-Ardakani, C. Jagger, T. Fujimori, L. Cholewa, V. Tilakaratna, J. Östling, M. Thomas, A.J. Day, R.J. Snelgrove, and T. Hussell. 2019. Defective lung function following influenza virus is due to prolonged, reversible hyaluronan synthesis. Matrix Biol. 80:14–28. doi:10.1016/j.matbio.2018.06.006.

3 Borthwick, L.A., L. Barron, K.M. Hart, K.M. Vannella, R.W. Thompson, S. Oland, A. Cheever, J. Sciurba, T.R. Ramalingam, A.J. Fisher, and T.A. Wynn. 2016. Macrophages are critical to the maintenance of IL-13-dependent lung inflammation and fibrosis. Mucosal Immunol. 9:38–55. doi:10.1038/mi.2015.34.

4 Cantuti-Castelvetri, L., R. Ojha, L.D. Pedro, M. Djannatian, J. Franz, S. Kuivanen, F. van der Meer, K. Kallio, T. Kaya, M. Anastasina, T. Smura, L. Levanov, L. Szirovicza, A. Tobi, H. Kallio-Kokko, P. Österlund, M. Joensuu, F.A. Meunier, S.J. Butcher, M.S. Winkler, B. Mollenhauer, A. Helenius, O. Gokce, T. Teesalu, J. Hepojoki, O. Vapalahti, C. Stadelmann, G. Balistreri, and M. Simons. 2020. Neuropilin-1 facilitates SARS-CoV-2 cell entry and infectivity. Science. 370:856–860. doi:10.1126/science.abd2985.

5 Chiaramonte, M. 2001. Studies of murine schistosomiasis reveal interleukin-13 blockade as a treatment for established and progressive liver fibrosis. Hepatology. 34:273–282. doi:10.1053/jhep.2001.26376.

6 Del Valle, D.M., S. Kim-Schulze, H.-H. Huang, N.D. Beckmann, S. Nirenberg, B. Wang, Y. Lavin, T.H. Swartz, D. Madduri, A. Stock, T.U. Marron, H. Xie, M. Patel, K. Tuballes, O. Van Oekelen, A. Rahman, P. Kovatch, J.A. Aberg, E. Schadt, S. Jagannath, M. Mazumdar, A.W. Charney, A. Firpo-Betancourt, D.R. Mendu, J. Jhang, D. Reich, K. Sigel, C. Cordon-Cardo, M. Feldmann, S. Parekh, M. Merad, and S. Gnjatic. 2020. An inflammatory cytokine signature predicts COVID-19 severity and survival. Nat. Med. 26:1636–1643. doi:10.1038/s41591-020-1051-9.

7 Ding, M., Q. Zhang, Q. Li, T. Wu, and Y. Huang. 2020. Correlation analysis of the severity and clinical prognosis of 32 cases of patients with COVID-19. Respir. Med. 167:105981. doi:10.1016/j.rmed.2020.105981.

8 Dobin, A., and T.R. Gingeras. 2015. Mapping RNA□seq Reads with STAR. Curr. Protoc. Bioinforma. 51. doi:10.1002/0471250953bi1114s51.

9 Ewels, P., M. Magnusson, S. Lundin, and M. Käller. 2016. MultiQC: summarize analysis results for multiple tools and samples in a single report. Bioinformatics. 32:3047–3048. doi:10.1093/bioinformatics/btw354.

10 Griss, J., G. Viteri, K. Sidiropoulos, V. Nguyen, A. Fabregat, and H. Hermjakob. 2020. ReactomeGSA - Efficient Multi-Omics Comparative Pathway Analysis. Mol. Cell. Proteomics. 19:2115–2125. doi:10.1074/mcp.TIR120.002155.

11 Heldin, P., C. Kolliopoulos, C.-Y. Lin, and C.-H. Heldin. 2020. Involvement of hyaluronan and CD44 in cancer and viral infections. Cell. Signal. 65:109427. doi:10.1016/j.cellsig.2019.109427.

12 Hellman, U., M.G. Karlsson, A. Engström-Laurent, S. Cajander, L. Dorofte, C. Ahlm, C. Laurent, and A. Blomberg. 2020. Presence of hyaluronan in lung alveoli in severe Covid-19: An opening for new treatment options? J. Biol. Chem. 295:15418–15422. doi:10.1074/jbc.AC120.015967.

13 Hoffmann, M., H. Kleine-Weber, S. Schroeder, N. Krüger, T. Herrler, S. Erichsen, T.S. Schiergens, G. Herrler, N.-H. Wu, A. Nitsche, M.A. Müller, C. Drosten, and S. Pöhlmann. 2020. SARS-CoV-2 Cell Entry Depends on ACE2 and TMPRSS2 and Is Blocked by a Clinically Proven Protease Inhibitor. Cell. 181:271–280.e8. doi:10.1016/j.cell.2020.02.052.

14 Honda, T., T. Kaneiwa, S. Mizumoto, K. Sugahara, and S. Yamada. 2012. Hyaluronidases Have Strong Hydrolytic Activity toward Chondroitin 4-Sulfate Comparable to that for Hyaluronan. Biomolecules. 2:549–563. doi:10.3390/biom2040549.

15 Huang, C., Y. Wang, X. Li, L. Ren, J. Zhao, Y. Hu, L. Zhang, G. Fan, J. Xu, X. Gu, Z. Cheng, T. Yu, J. Xia, Y. Wei, W. Wu, X. Xie, W. Yin, H. Li, M. Liu, Y. Xiao, H. Gao, L. Guo, J. Xie, G. Wang, R. Jiang, Z. Gao, Q. Jin, J. Wang, and B. Cao. 2020. Clinical features of patients infected with 2019 novel coronavirus in Wuhan, China. The Lancet. 395:497–506. doi:10.1016/S0140-6736(20)30183-5.

16 Kalil, A.C., T.F. Patterson, A.K. Mehta, K.M. Tomashek, C.R. Wolfe, V. Ghazaryan, V.C. Marconi, G.M. Ruiz-Palacios, L. Hsieh, S. Kline, V. Tapson, N.M. Iovine, M.K. Jain, D.A. Sweeney, H.M. El Sahly, A.R. Branche, J. Regalado Pineda, D.C. Lye, U. Sandkovsky, A.F. Luetkemeyer, S.H. Cohen, R.W. Finberg, P.E.H. Jackson, B. Taiwo, C.I. Paules, H. Arguinchona, N. Erdmann, N. Ahuja, M. Frank, M. Oh, E.-S. Kim, S.Y. Tan, R.A. Mularski, H. Nielsen, P.O. Ponce, B.S. Taylor, L. Larson, N.G. Rouphael, Y. Saklawi, V.D. Cantos, E.R. Ko, J.J. Engemann, A.N. Amin, M. Watanabe, J. Billings, M.-C. Elie, R.T. Davey, T.H. Burgess, J. Ferreira, M. Green, M. Makowski, A. Cardoso, S. de Bono, T. Bonnett, M. Proschan, G.A. Deye, W. Dempsey, S.U. Nayak, L.E. Dodd, and J.H. Beigel. 2020. Baricitinib plus Remdesivir for Hospitalized Adults with Covid-19. N. Engl. J. Med. NEJMoa2031994. doi:10.1056/NEJMoa2031994.

17 Lauer, M.E., R.A. Dweik, S. Garantziotis, and M.A. Aronica. 2015. The Rise and Fall of Hyaluronan in Respiratory Diseases. Int. J. Cell Biol. 2015:1–15. doi:10.1155/2015/712507.

18 Le Floc’h, A., J. Allinne, K. Nagashima, G. Scott, D. Birchard, S. Asrat, Y. Bai, W.K. Lim, J. Martin, T. Huang, T.B. Potocky, J.H. Kim, A. Rafique, N.J. Papadopoulos, N. Stahl, G.D. Yancopoulos, A.J. Murphy, M.A. Sleeman, and J.M. Orengo. 2020. Dual blockade of IL□4 and IL□13 with dupilumab, an IL□4R*α* antibody, is required to broadly inhibit type 2 inflammation. Allergy. 75:1188–1204. doi:10.1111/all.14151.

19 Lee, C.G., C.A. Da Silva, C.S. Dela Cruz, F. Ahangari, B. Ma, M.-J. Kang, C.-H. He, S. Takyar, and J.A. Elias. 2011. Role of Chitin and Chitinase/Chitinase-Like Proteins in Inflammation, Tissue Remodeling, and Injury. Annu. Rev. Physiol. 73:479–501. doi:10.1146/annurev-physiol-012110-142250.

20 Liao, M., Y. Liu, J. Yuan, Y. Wen, G. Xu, J. Zhao, L. Cheng, J. Li, X. Wang, F. Wang, L. Liu, I. Amit, S. Zhang, and Z. Zhang. 2020. Single-cell landscape of bronchoalveolar immune cells in patients with COVID-19. Nat. Med. 26:842–844. doi:10.1038/s41591-020-0901-9.

21 Liu, F., L. Li, M. Xu, J. Wu, D. Luo, Y. Zhu, B. Li, X. Song, and X. Zhou. 2020. Prognostic value of interleukin-6, C-reactive protein, and procalcitonin in patients with COVID-19. J. Clin. Virol. 127:104370. doi:10.1016/j.jcv.2020.104370.

22 Love, M.I., W. Huber, and S. Anders. 2014. Moderated estimation of fold change and dispersion for RNA-seq data with DESeq2. Genome Biol. 15:550. doi:10.1186/s13059-014-0550-8.

23 Lucas, C., P. Wong, J. Klein, T. Castro, J. Silva, M. Sundaram, M. Ellingson, T. Mao, J. Oh, B. Israelow, M. Tokuyama, P. Lu, A. Venkataraman, A. Park, S. Mohanty, H. Wang, A.L. Wyllie, C.B.F. Vogels, R. Earnest, S. Lapidus, I. Ott, A. Moore, C. Muenker, J. Fournier, M. Campbell, C. Odio, A. Casanovas-Massana, Yale IMPACT Team, R. Herbst, A. Shaw, R. Medzhitov, W.L. Schulz, N. Grubaugh, C. Dela Cruz, S. Farhadian, A. Ko, S. Omer, and A. Iwasaki. 2020. Longitudinal immunological analyses reveal inflammatory misfiring in severe COVID-19 patients. Infectious Diseases (except HIV/AIDS).

24 Mangalmurti, N., and C.A. Hunter. 2020. Cytokine Storms: Understanding COVID-19. Immunity. 53:19–25. doi:10.1016/j.immuni.2020.06.017.

25 Marone, G., F. Granata, V. Pucino, A. Pecoraro, E. Heffler, S. Loffredo, G.W. Scadding, and G. Varricchi. 2019. The Intriguing Role of Interleukin 13 in the Pathophysiology of Asthma. Front. Pharmacol. 10:1387. doi:10.3389/fphar.2019.01387.

26 Martin, M. 2011. Cutadapt removes adapter sequences from high-throughput sequencing reads. EMBnet.journal. 17:10. doi:10.14806/ej.17.1.200.

27 Matute-Bello, G., G. Downey, B.B. Moore, S.D. Groshong, M.A. Matthay, A.S. Slutsky, and W.M. Kuebler. 2011. An Official American Thoracic Society Workshop Report: Features and Measurements of Experimental Acute Lung Injury in Animals. Am. J. Respir. Cell Mol. Biol. 44:725–738. doi:10.1165/rcmb.2009-0210ST.

28 McCormick, S.M., and N.M. Heller. 2015. Commentary: IL-4 and IL-13 receptors and signaling. Cytokine. 75:38–50. doi:10.1016/j.cyto.2015.05.023.

29 Misra, S., V.C. Hascall, R.R. Markwald, and S. Ghatak. 2015. Interactions between Hyaluronan and Its Receptors (CD44, RHAMM) Regulate the Activities of Inflammation and Cancer. Front. Immunol. 6. doi:10.3389/fimmu.2015.00201.

30 Moreau, G.B., S.L. Burgess, J.M. Sturek, A.N. Donlan, W.A. Petri, and B.J. Mann. 2020. Evaluation of K18-hACE2 Mice as a Model of SARS-CoV-2 Infection. Am. J. Trop. Med. Hyg. 103:1215–1219. doi:10.4269/ajtmh.20-0762.

31 Nagy, N., H.F. Kuipers, P.L. Marshall, E. Wang, G. Kaber, and P.L. Bollyky. 2019. Hyaluronan in immune dysregulation and autoimmune diseases. Matrix Biol. 78–79:292– 313. doi:10.1016/j.matbio.2018.03.022.

32 Nienhold, R., Y. Ciani, V.H. Koelzer, A. Tzankov, J.D. Haslbauer, T. Menter, N. Schwab, M. Henkel, A. Frank, V. Zsikla, N. Willi, W. Kempf, T. Hoyler, M. Barbareschi, H. Moch, M. Tolnay, G. Cathomas, F. Demichelis, T. Junt, and K.D. Mertz. 2020. Two distinct immunopathological profiles in autopsy lungs of COVID-19. Nat. Commun. 11:5086. doi:10.1038/s41467-020-18854-2.

33 Pedersen, S.F., and Y.-C. Ho. 2020. SARS-CoV-2: a storm is raging. J. Clin. Invest. 130:2202–2205. doi:10.1172/JCI137647.

34 Petrey, A.C., F. Qeadan, E.A. Middleton, I.V. Pinchuk, R.A. Campbell, and E.J. Beswick. 2020. Cytokine release syndrome in COVID-19: Innate immune, vascular, and platelet pathogenic factors differ in severity of disease and sex. J. Leukoc. Biol. doi:10.1002/JLB.3COVA0820-410RRR.

35 Rathnasinghe, R., S. Strohmeier, F. Amanat, V.L. Gillespie, F. Krammer, A. García-Sastre, L. Coughlan, M. Schotsaert, and M.B. Uccellini. 2020. Comparison of transgenic and adenovirus hACE2 mouse models for SARS-CoV-2 infection. Emerg. Microbes Infect. 9:2433–2445. doi:10.1080/22221751.2020.1838955.

36 Rayahin, J.E., J.S. Buhrman, Y. Zhang, T.J. Koh, and R.A. Gemeinhart. 2015. High and Low Molecular Weight Hyaluronic Acid Differentially Influence Macrophage Activation. ACS Biomater. Sci. Eng. 1:481–493. doi:10.1021/acsbiomaterials.5b00181.

37 Ruppert, S.M., T.R. Hawn, A. Arrigoni, T.N. Wight, and P.L. Bollyky. 2014. Tissue integrity signals communicated by high-molecular weight hyaluronan and the resolution of inflammation. Immunol. Res. 58:186–192. doi:10.1007/s12026-014-8495-2.

38 Su, H., M. Yang, C. Wan, L.-X. Yi, F. Tang, H.-Y. Zhu, F. Yi, H.-C. Yang, A.B. Fogo, X. Nie, and C. Zhang. 2020. Renal histopathological analysis of 26 postmortem findings of patients with COVID-19 in China. Kidney Int. 98:219–227. doi:10.1016/j.kint.2020.04.003.

39 The RECOVERY Collaborative Group. 2020. Dexamethasone in Hospitalized Patients with Covid-19 — Preliminary Report. N. Engl. J. Med. NEJMoa2021436. doi:10.1056/NEJMoa2021436.

40 Tisoncik, J.R., M.J. Korth, C.P. Simmons, J. Farrar, T.R. Martin, and M.G. Katze. 2012. Into the Eye of the Cytokine Storm. Microbiol. Mol. Biol. Rev. 76:16–32. doi:10.1128/MMBR.05015-11.

41 Weiskopf, D., K.S. Schmitz, M.P. Raadsen, A. Grifoni, N.M.A. Okba, H. Endeman, J.P.C. van den Akker, R. Molenkamp, M.P.G. Koopmans, E.C.M. van Gorp, B.L. Haagmans, R.L. de Swart, A. Sette, and R.D. de Vries. 2020. Phenotype and kinetics of SARS-CoV-2-specific T cells in COVID-19 patients with acute respiratory distress syndrome. Sci. Immunol. 5:eabd2071. doi:10.1126/sciimmunol.abd2071.

42 Winkler, E.S., A.L. Bailey, N.M. Kafai, S. Nair, B.T. McCune, J. Yu, J.M. Fox, R.E. Chen, J.T. Earnest, S.P. Keeler, J.H. Ritter, L.-I. Kang, S. Dort, A. Robichaud, R. Head, M.J. Holtzman, and M.S. Diamond. 2020. Publisher Correction: SARS-CoV-2 infection of human ACE2-transgenic mice causes severe lung inflammation and impaired function. Nat. Immunol. 21:1470–1470. doi:10.1038/s41590-020-0794-2.

43 Wu, D., and G.K. Smyth. 2012. Camera: a competitive gene set test accounting for inter-gene correlation. Nucleic Acids Res. 40:e133–e133. doi:10.1093/nar/gks461.

44 Yang, Y., C. Shen, J. Li, J. Yuan, J. Wei, F. Huang, F. Wang, G. Li, Y. Li, L. Xing, L. Peng, M. Yang, M. Cao, H. Zheng, W. Wu, R. Zou, D. Li, Z. Xu, H. Wang, M. Zhang, Z. Zhang, G.F. Gao, C. Jiang, L. Liu, and Y. Liu. 2020. Plasma IP-10 and MCP-3 levels are highly associated with disease severity and predict the progression of COVID-19. J. Allergy Clin. Immunol. 146:119–127.e4. doi:10.1016/j.jaci.2020.04.027.

