## Supplemental Materials for "IL-13 is a driver of COVID-19 severity"

**Title: IL-13 is a driver of COVID-19 severity**

Fig. S1. Visualization of cytokines in outpatients and inpatients with COVID-19 and in uninfected controls. A) A total of 47 cytokines, chemokines and growth factors were measured from plasma of COVID-19 positive patients, and cytokines of interest were compared between patients with differing severity of illness using a Mann-Whitney U test. (OP, outpatient; IP, inpatient; IP+Vent, ventilated inpatient). B) 26 cytokines and growth factors were measured in plasma from 19 non-severe and 26 severe (requiring supplemental oxygen) COVID patients from VCU. Cytokines of interest were compared using a Mann-Whitney U test.

Fig. S2. Network analysis of cytokine genes in patients with COVID-19. The network analysis captured the structural relationships among cytokine measurements with graphical LASSO. The nodes represent individual cytokines and edges represent their correlations. Highly correlated cytokines are connected closer with thick edges. Green line = positive correlation; red line = negative correlation. Key indicates abbreviations used for each cytokine. B) Linear regression between plasma IL-13 levels and time from symptom onset to blood draw. Blue = did not require ventilation; Red = required ventilation.

**Fig. S3. Impact of anti-IL-13 on lung injury and inflammation in a mouse model of COVID-19.** Mice were infected with 5x10^3^ PFU of SARS-CoV-2 on day 0 and given 150 µg of anti-IL-13 or IgG isotype control on days 0, 2, and 4. On day five, mice were euthanized and bronchoalveolar lavage (BAL) fluid collected. For histology, lungs were inflated with formalin before removing and fixing prior to H&E staining. A) Viral burden in lungs on day five pi was measured by plaque forming units (PFU). B) Lung injury score of infected mouse lung with or without anti-IL-13. C) Hematoxylin and eosin stain. D) Masson’s trichrome staining of lungs on day 8. E) Modified Ashcroft score. Upper panel scale bar = 1.5 mm. Lower panel scale bars = 90 µm. F) Ym1 with or without anti-IL-13. G) Cytokines in BAL measured by Luminex (plotted with group identity and clinical score). H) immune cells in BAL quantified by flow cytometry.

**Fig. S4. Contributions of hyaluronan and its receptor to severe COVID-19 in the mouse model.** Mice were infected with 5x10^3^ PFU of SARS-CoV-2 on day 0 and given 150 µg of anti-IL-13 or IgG isotype control on days 0, 2, and 4. On day five, mice were euthanized and sections of lung were stored in trizol. RNAseq was done on lung tissue. Read counts of A) hyaluronan synthase 2 (*Has2*) and B) hyaluronidases 1 and 2 (*Hyal1* and *Hyal2*) were analyzed between anti-IL-13 treated mice and isotype controls. C) Quantification of intensity of epithelial hyaluronan from fluorescent staining (log-transformed, mixed-model). D) Kaplan-Meier survival curve, E) clinical scores and F) weight loss from mice infected with 5x10^3^ PFU of SARS-CoV-2 who were administered hyaluronidase on day five pi; combined, two independent experiments. Infected mice were administered anti-CD44 antibodies or isotype IgG2 control on days 1, 2, 3 and 4 pi G) weight loss was quantified; data are combined from two, independent experiments. H) Lung sections stained with hyaluronan binding protein (HABP, green) and DAPI nuclei stain (blue) for SARS-CoV-2 infected mice treated with IgG or anti-IL-13. Each image is representative of an individual; scale bar = 70um.¨ *=p<0.05; **=p<0.005

**Fig. S5. Staining of Ym1, RELMα and HA for all experimental mice.** (Related to Fig. 3 and 4). Mice received i.p. injections of anti-IL-13 on days 0 and 2 pi, were euthanized on day 5. A) Lung sections stained with Ym1 (red) and RELMα (yellow) for SARS-CoV-2 infected mice treated with IgG or anti-IL-13. Each image is representative of an individual mouse, scale bar = 70µm. B) Lung sections stained with hyaluronan binding protein (HABP, green) and DAPI nuclei stain (blue) for SARS-CoV-2 infected mice treated with IgG or anti-IL-13. Each image is representative of an individual mouse, scale bar = 70µm

Table S1. Age, sex and clinical status of UVA patients.

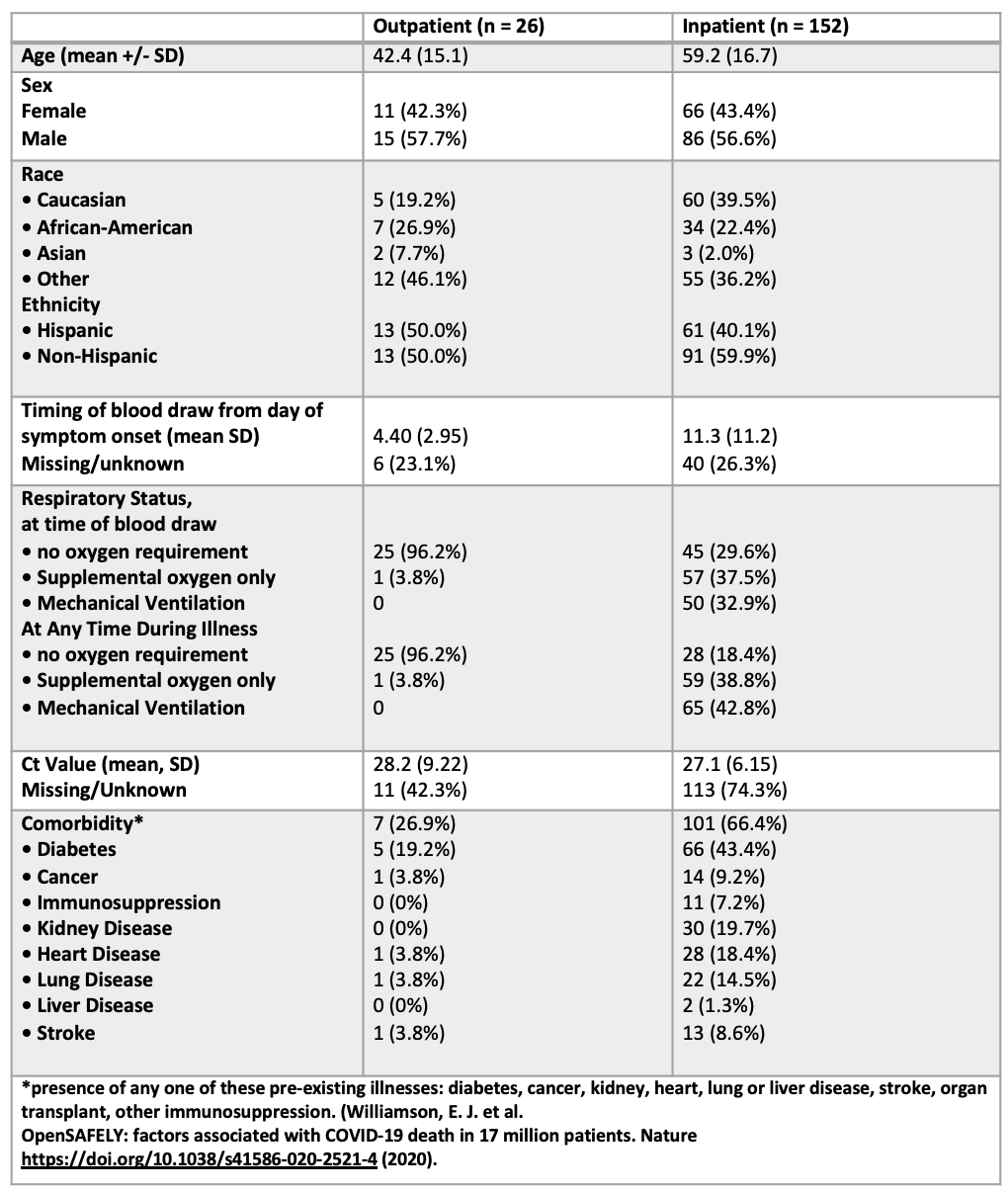

**Table S2. Contributors to Component 1 in PCA plot.** Principal component analysis was performed using the Proc Factor in SAS. For principal component one, those cytokines with a loading score of 0.5 or above were retained.

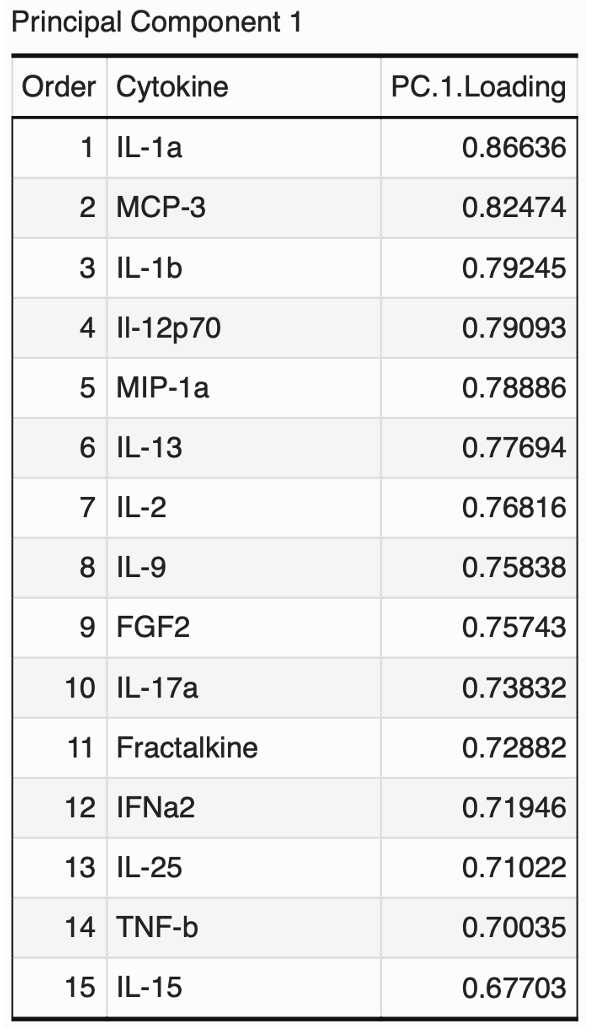

Table S3. Age, sex and clinical status of VCU patients.
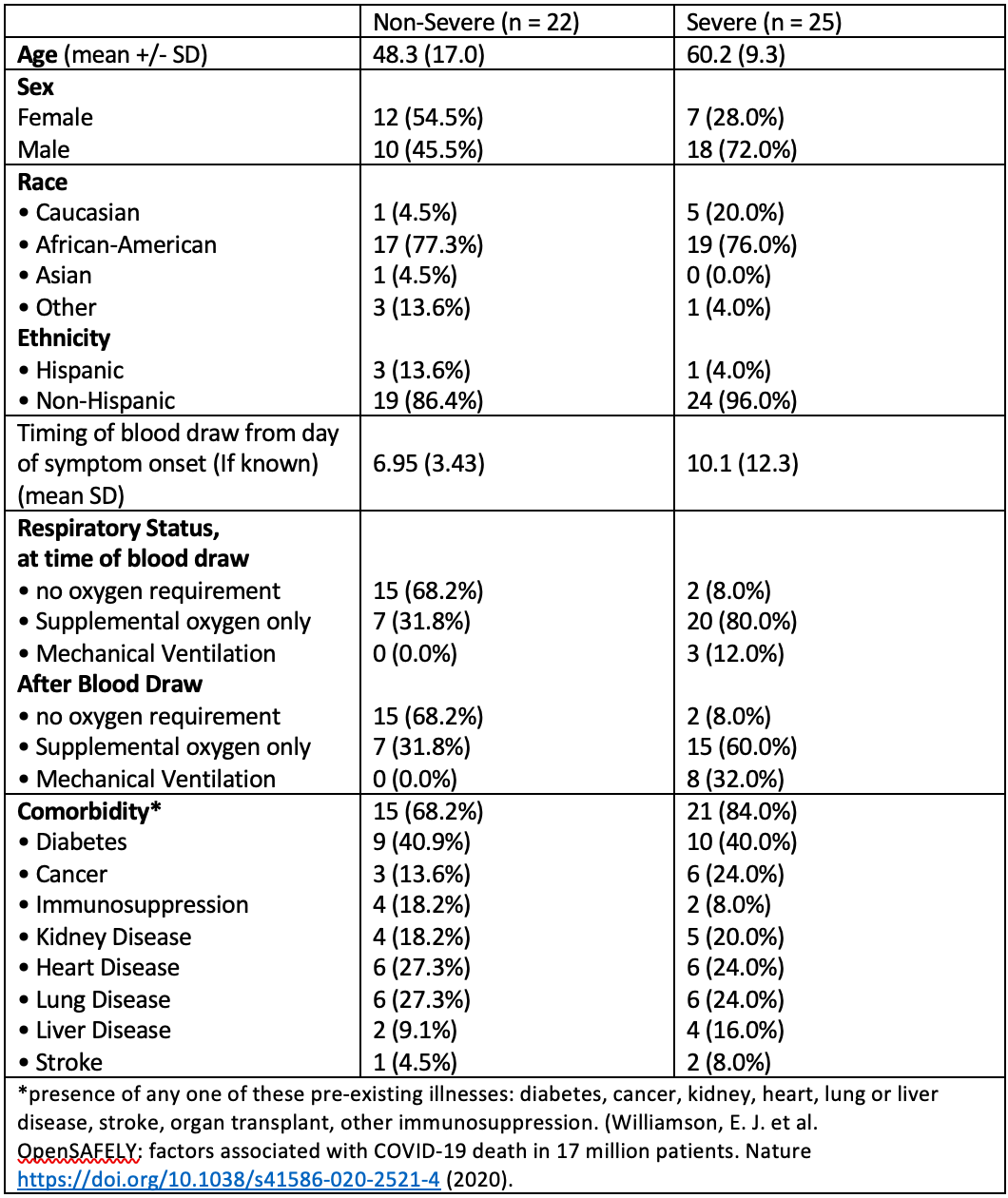

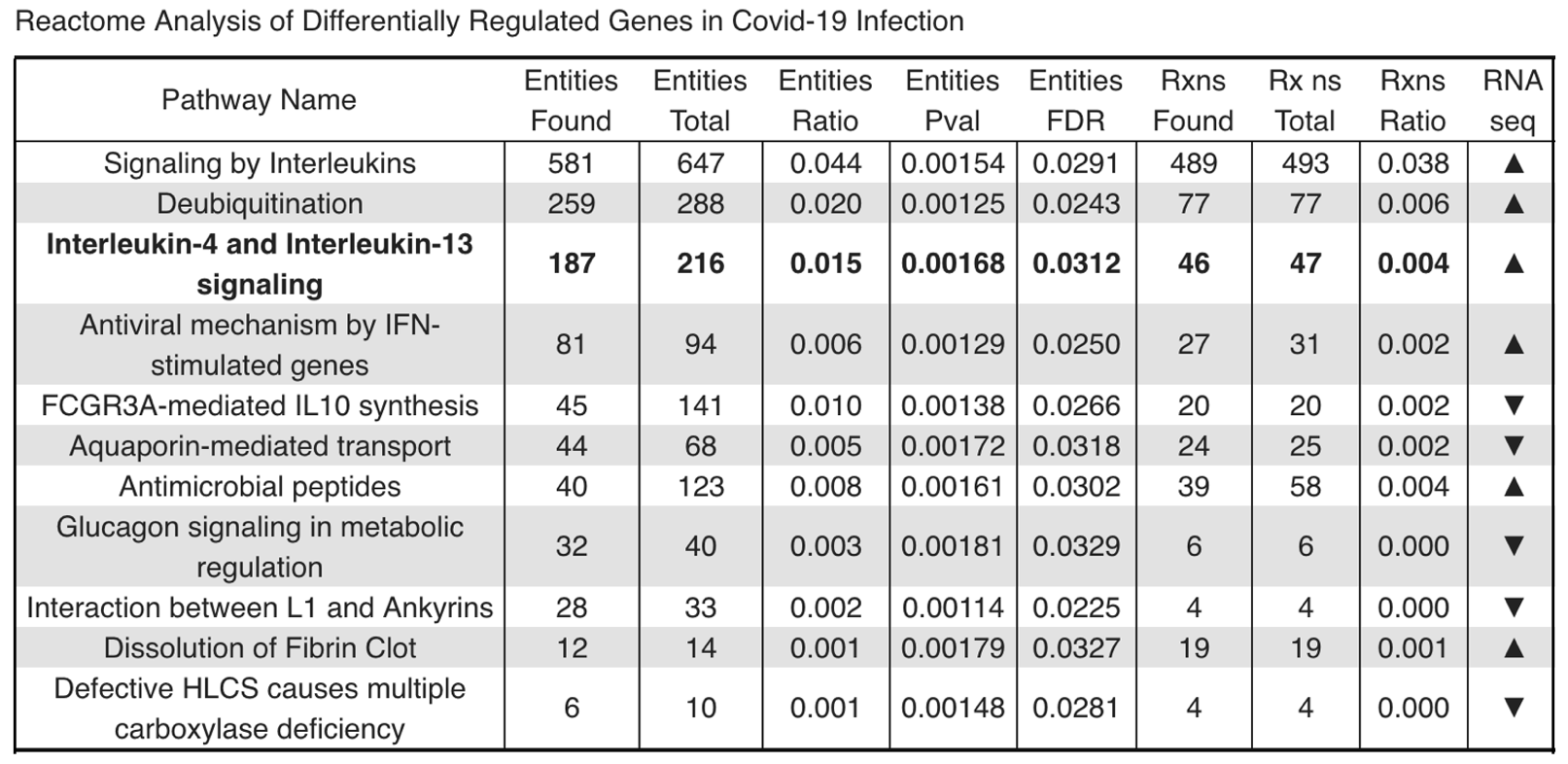
Table S4. Enriched pathways in differentially regulated genes in murine COVID-19 infection. Enrichment analysis was applied to the total gene counts using the CAMERA algorithm. Pathways are arranged by descending number of entities found.

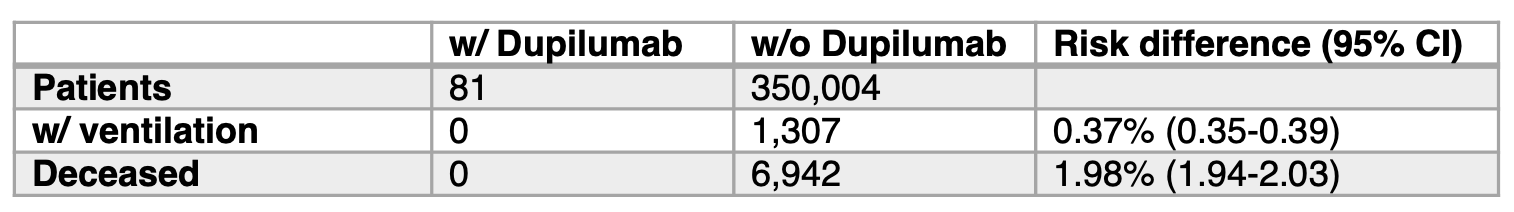
**Table S5. Outcomes with and without Dupilumab for the full COVID-19 cohort.**

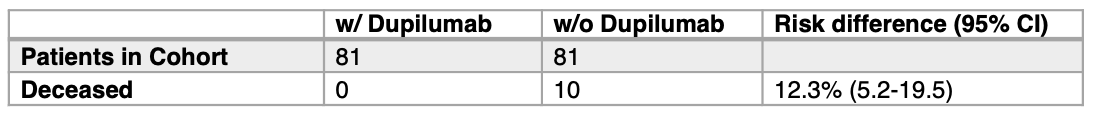
**Table S6. Sub cohort using 1:1 propensity score matching.**

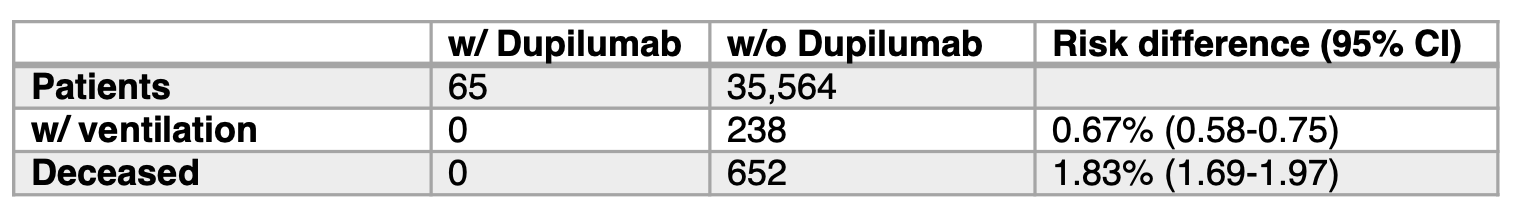
**Table S7. A sub cohort with diagnoses of either asthma, atopic dermatitis or rhinosinusitis.**

**Table S8. Lab values and standard deviations (SD) for serum levels of C-reactive protein (CRP) for COVID-19 patients in this study**

| **Group** | **CRP (mg/l)** | **SD** | **Sub-cohort difference** |
| --- | --- | --- | --- |
| **Deceased** | **100.0** | **107.0** | **65.7** |
| **Survived** | **34.3** | **58.0** |  |
| **Ventilated** | **74.9** | **98.5** | **35.3** |
| **Not Ventilated** | **39.6** | **65.6** |  |
| **Without Dupilumab** | **37.6** | **63.8** | **12.3** |
| **With Dupilumab** | **25.3** | **68.3** |  |

**Table S9.** **N3C Dupilumab COVID severity outcomes**. A total of 785 patients with a record of dupilumab prescription were in the N3C Data Enclave on November 16, 2020. Of these, 31 Dupilumab patients had a COVID+ test within 61 days after a dupilumab dose, resulting in a test positivity rate of 3.9% (95% CI: 2.8, 5.6). A total of 247,391 COVID+ patients with no record of Dupilumab were available for selection of matched controls. Five matches could be found for each patient. In the matched analytic dataset of COVID+ patients, no differences were seen in the hospitalization (OR=0.64, p=0.57) or death rates (p>0.99); though <20 deaths were seen in the entire dataset. In hospitalized patients, no differences were observed in the rates of ECMO (p>0.99) or IMV (p>0.99).

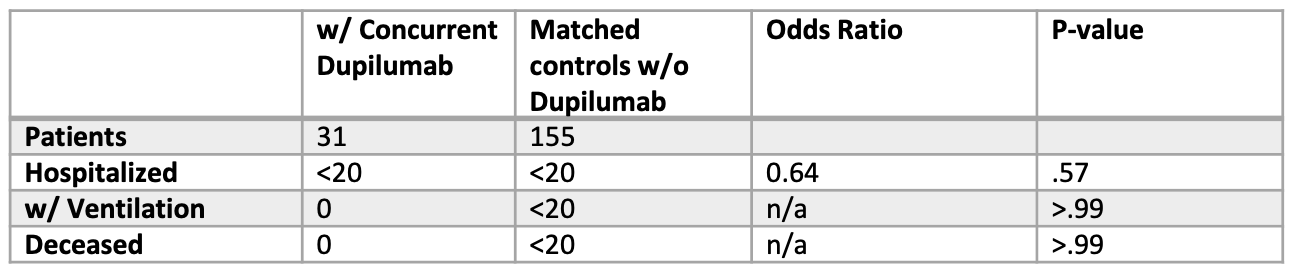

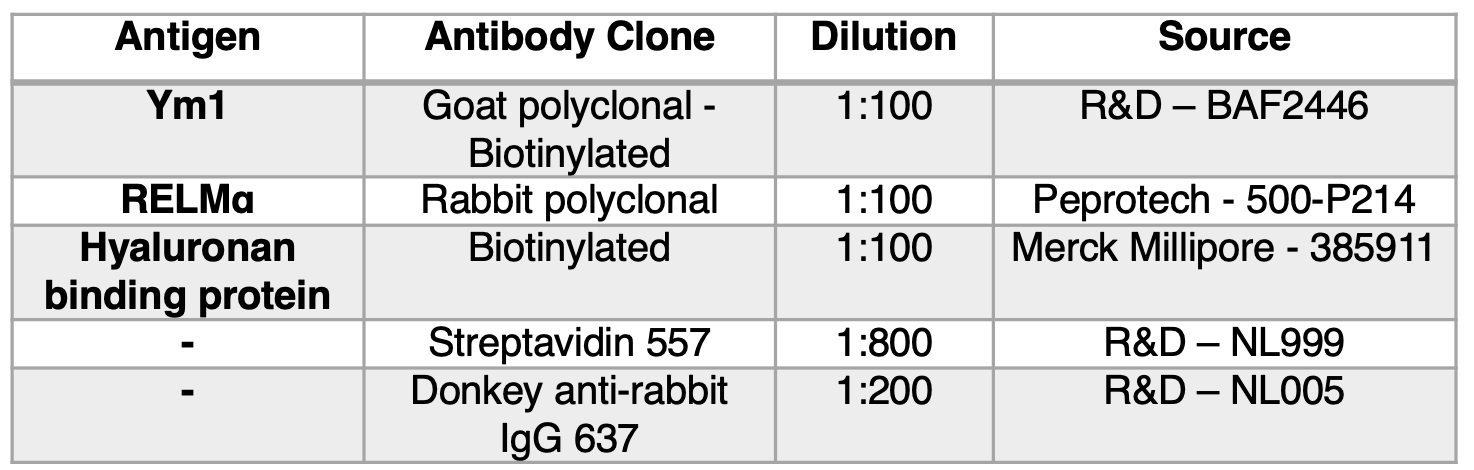
**Table S10. Immunohistochemistry reagents.**
