## Supplementary figures and images for "IL-13 is a driver of COVID-19 severity"

### Figure S1

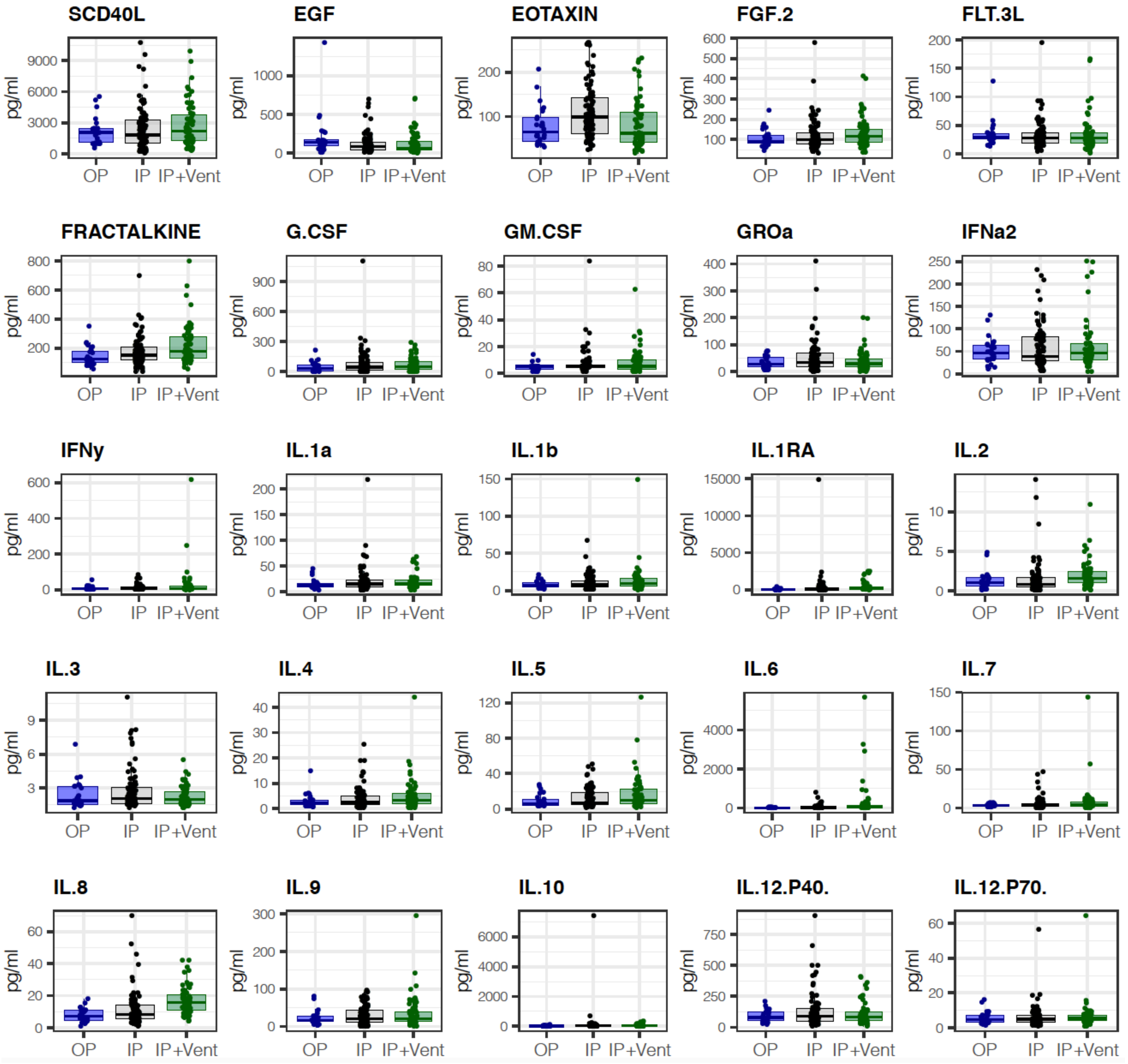

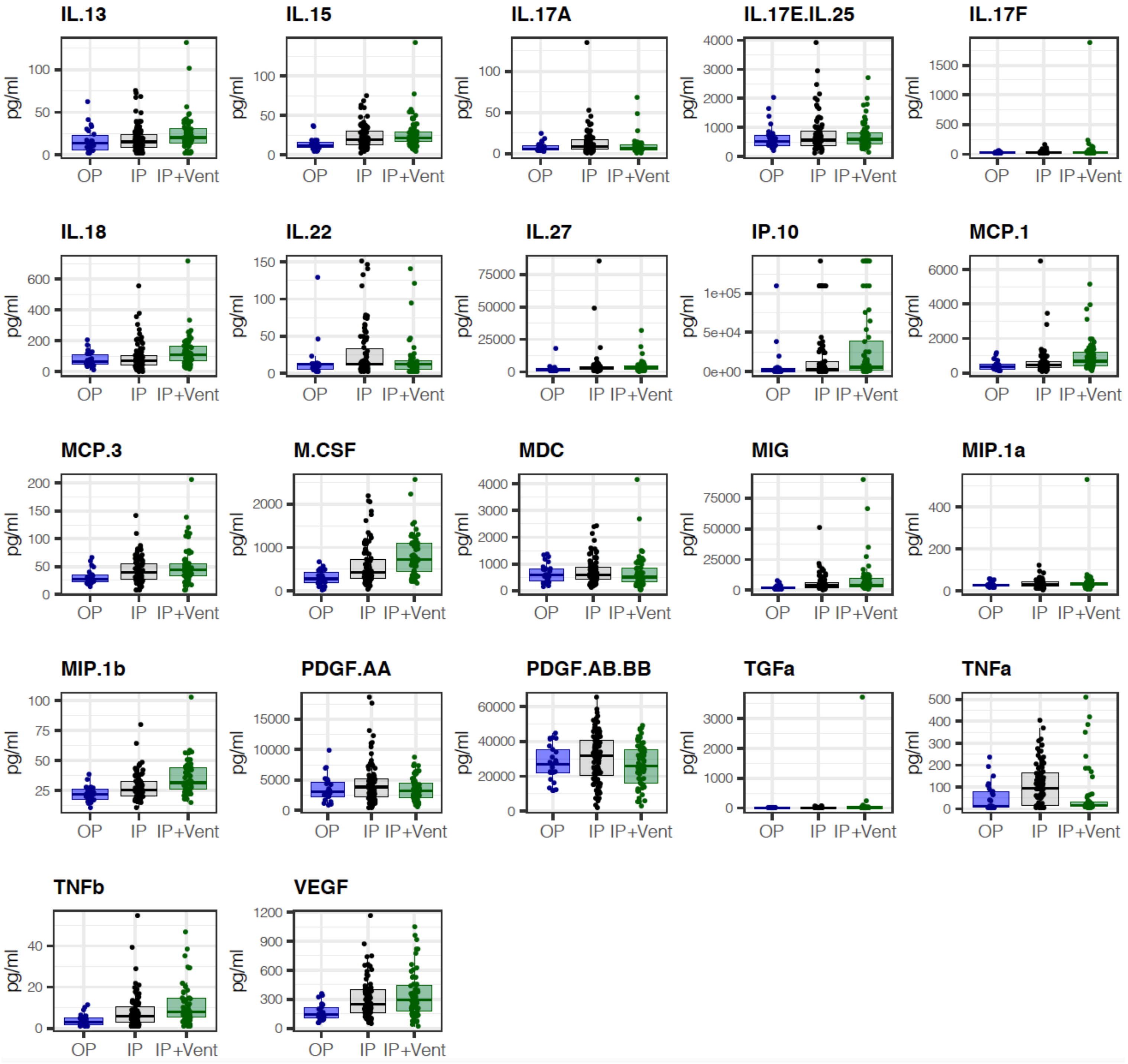

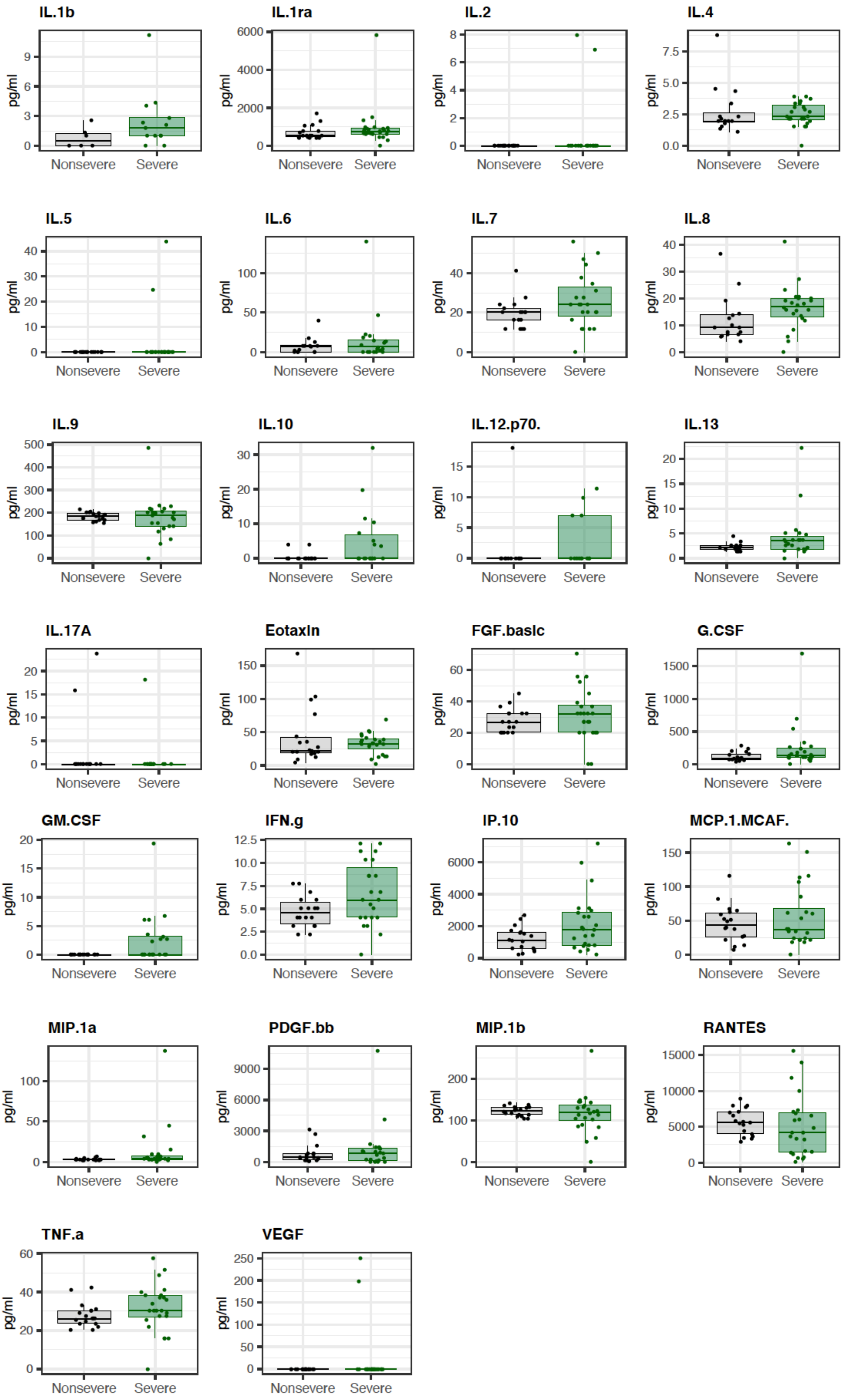

### Figure S2

Supp. Figure 2

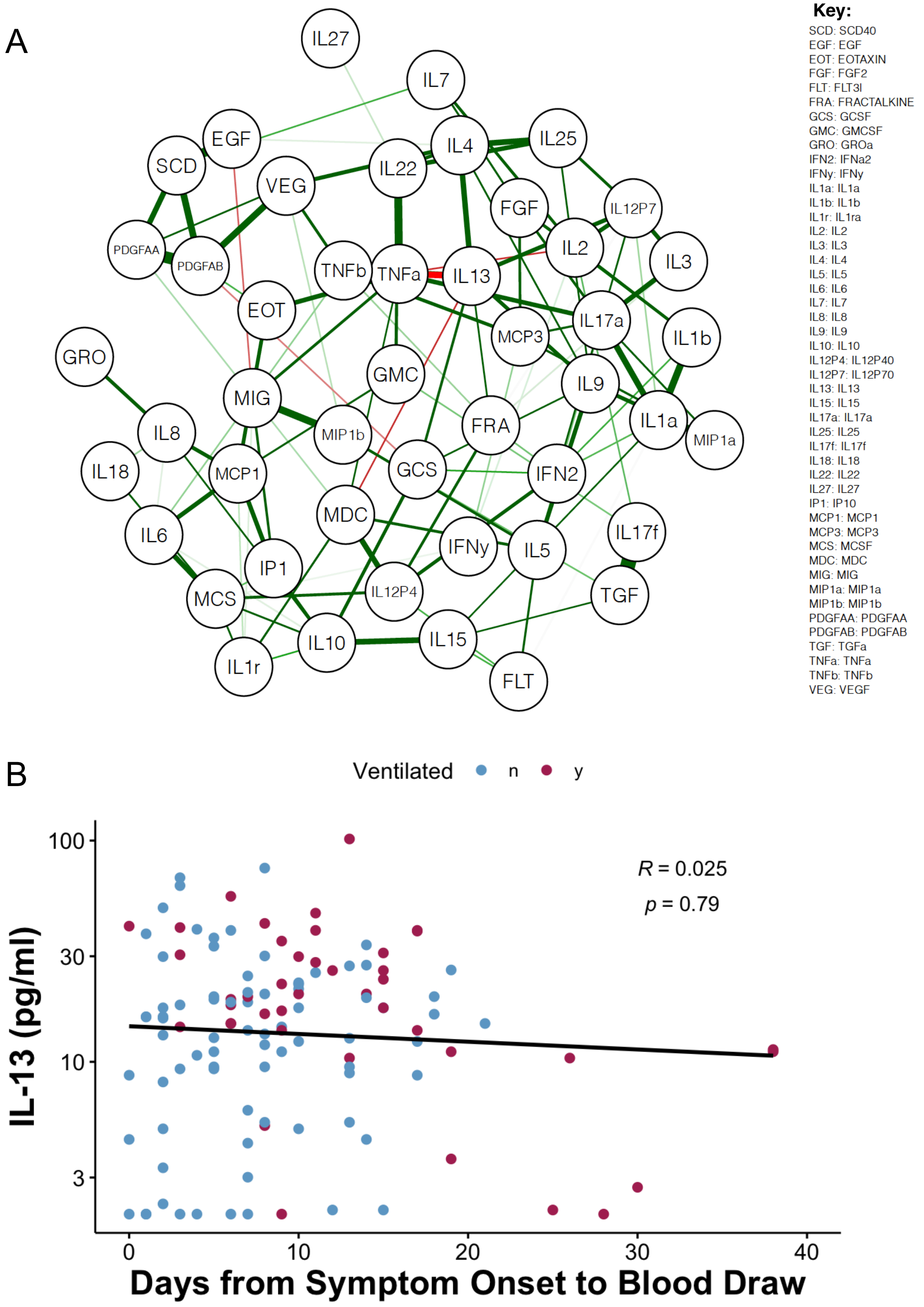

### Figure S3

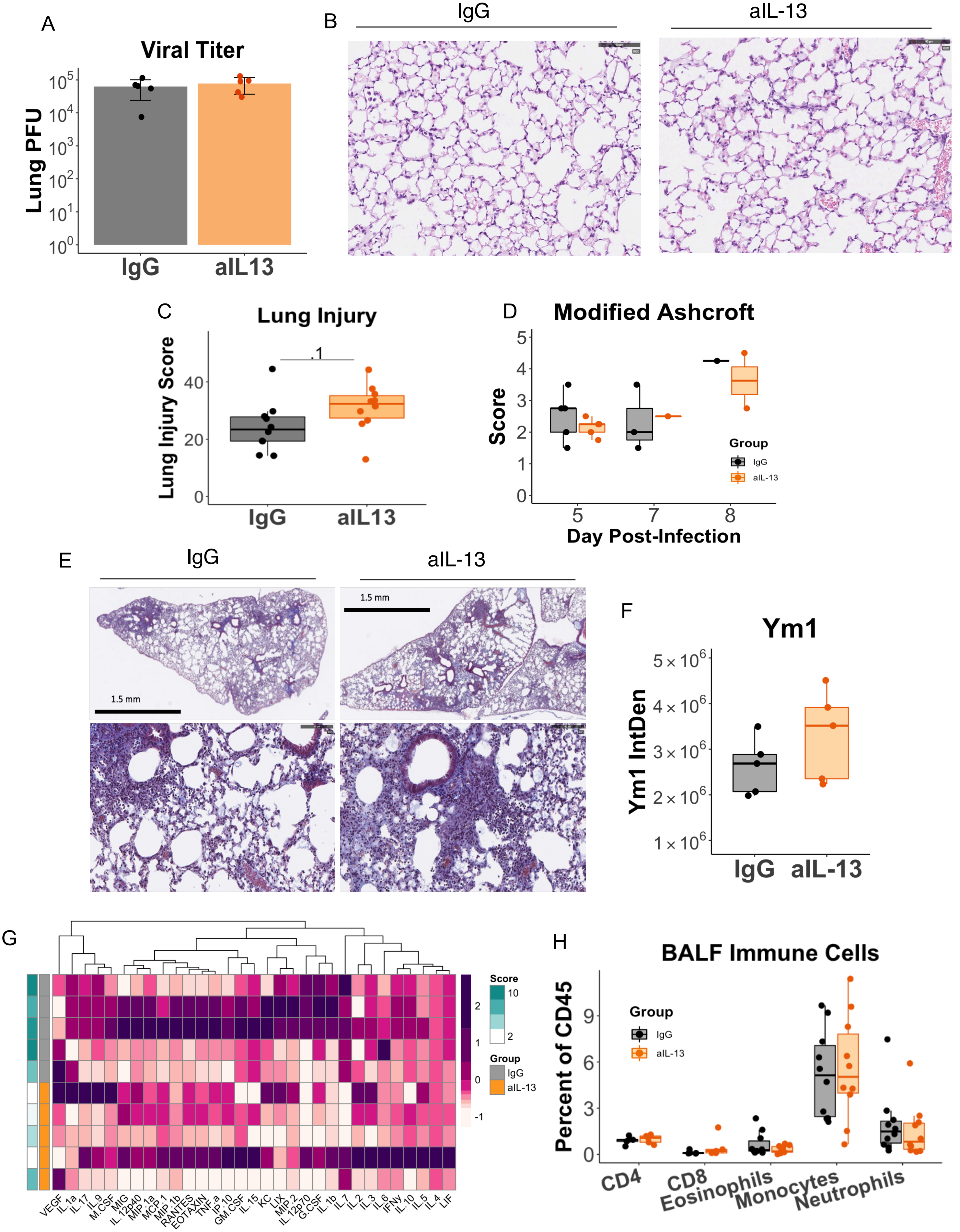

### Figure S4

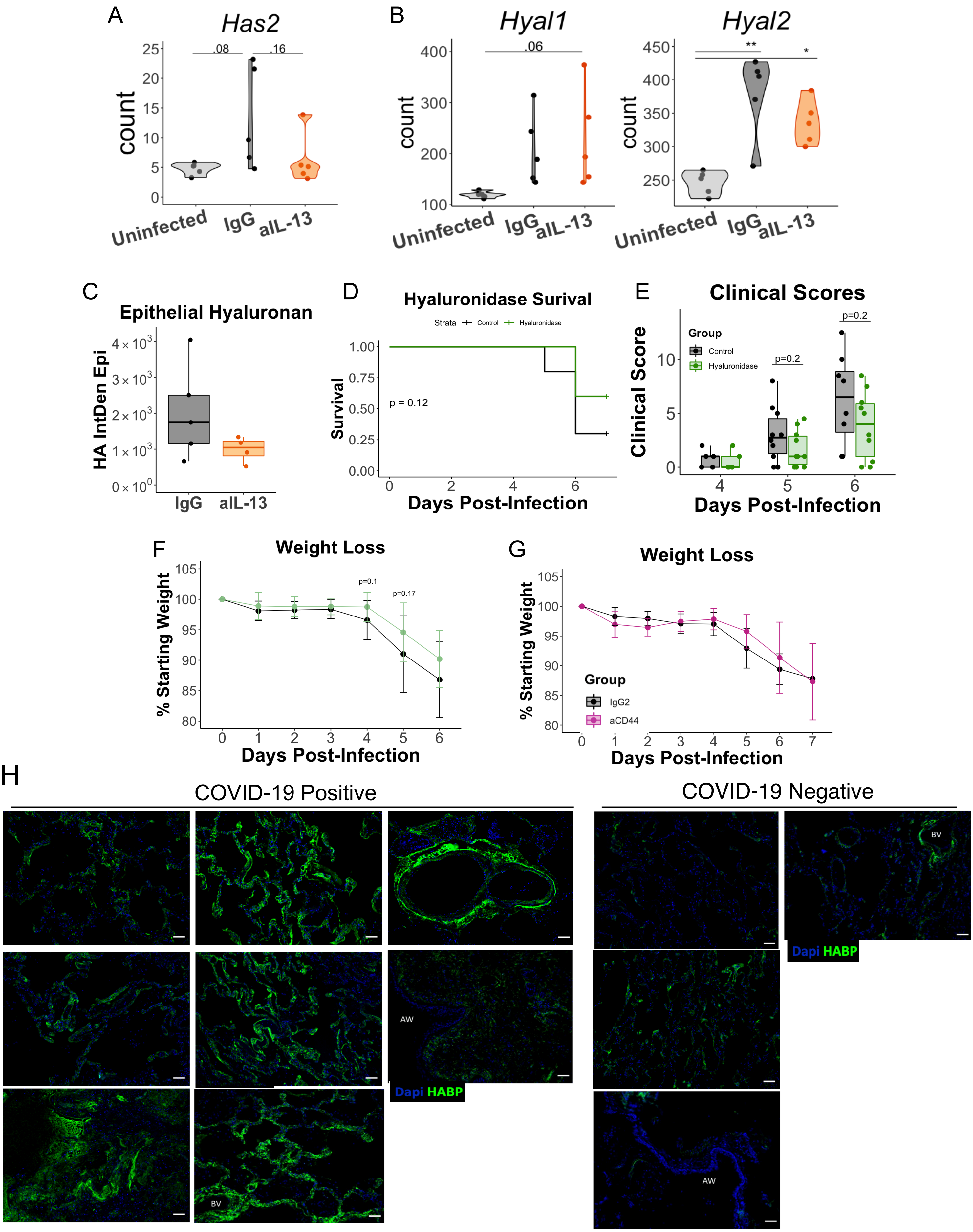

### Figure S5

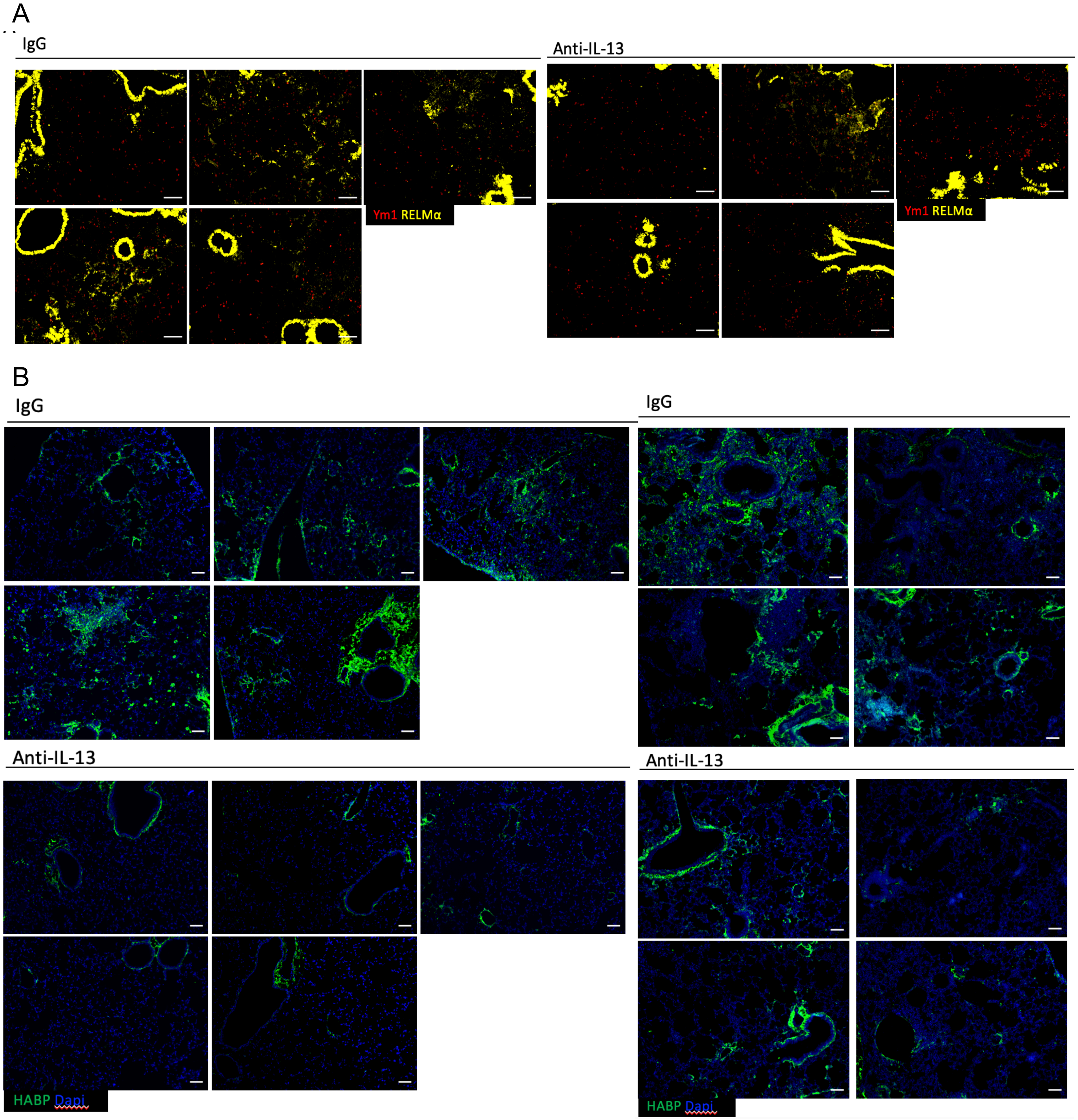
